# Enhanced EBNA2-dependent activity in EBV-transformed B cells from patients with multiple sclerosis

**DOI:** 10.64898/2026.02.18.26346386

**Authors:** Marissa Granitto, Ellie Kim, Carmy Forney, Cailing Yin, Arame A. Diouf, Andrew vonHandorf, Phillip Dexheimer, Sreeja Parameswaran, Xiaoting Chen, Omer Donmez, Hope Rowden, Casey Swoboda, Molly Shook, Katelyn Dunn, Hania Kebir, Maria Vélez-Colón, Kenneth Kaufman, Danielle Ho, Viktoryia Laurynenka, Lee E. Edsall, Veronica Brennan, Ben E. Gewurz, Bahram Namjou, Elizabeth Wilson, Kristen S. Fisher, Aram Zabeti, Lucinda P. Lawson, Jorge I. Alvarez, Leah C. Kottyan, Matthew T. Weirauch

**Affiliations:** Development, Stem Cells & Regenerative Medicine Graduate Program, University of Cincinnati College of Medicine, Cincinnati, OH, 45229, USA; Center for Autoimmune Genomics and Etiology, Cincinnati Children’s Hospital Medical Center, Cincinnati, OH, 45229, USA; Division of Allergy and Immunology, Cincinnati Children’s Hospital Medical Center, Cincinnati, OH, 45229, USA; Immunology Graduate Program, University of Cincinnati College of Medicine, Cincinnati, OH, 45229, USA; Medical Scientist Training Program, University of Cincinnati College of Medicine, Cincinnati, OH, 45229, USA; Division of Biomedical Informatics, Cincinnati Children’s Hospital Medical Center, Cincinnati, OH, 45229, USA; Department of Pathobiology, School of Veterinary Medicine, University of Pennsylvania, Philadelphia, PA, 19104, USA; Division of Human Genetics, Cincinnati Children’s Hospital Medical Center, Cincinnati, OH, 45229, USA; Research Service, US Department of Veterans Affairs Medical Center, Cincinnati, Ohio, USA; Department of Pediatrics, University of Cincinnati College of Medicine, Cincinnati, OH, 45229, USA; Department of Medicine, Division of Infectious Diseases, Brigham and Women’s Hospital, Harvard Medical School, Boston, MA, 02115, USA; Department of Microbiology, Harvard Program in Virology, Harvard Medical School, Boston, MA, 02115, USA; Center for Integrated Solutions to Infectious Diseases, Broad Institute of Harvard and MIT, Cambridge, MA, 02142, USA; Division of Neurology, Cincinnati Children’s Hospital Medical Center, Cincinnati, OH, 45229, USA; Department of Pediatrics, Division of Neurology and Developmental Neuroscience, Baylor College of Medicine, Houston, TX 77030, USA; Waddell Center for Multiple Sclerosis, College of Medicine, University of Cincinnati, Cincinnati, OH 45220, USA; Division of Developmental Biology, Cincinnati Children’s Hospital Medical Center, Cincinnati, OH, 45229, USA

## Abstract

Multiple sclerosis (MS) is an immune-mediated demyelinating disease of the central nervous system affecting 2.8 million people worldwide. Both genetic and environmental factors contribute to MS risk, with Epstein-Barr virus (EBV) infection being an important environmental factor. To better clarify the role of EBV in MS, we examined its impact on gene expression, chromatin accessibility, and transcription factor binding in primary B cells and EBV-transformed B cells derived from patients with MS and healthy controls. RNA-seq and ATAC-seq analyses revealed extensive MS-dependent gene expression and chromatin accessibility differences in EBV-transformed, but not in primary B cells. These changes are largely accounted for by the expression levels of EBNA2, an EBV-encoded transcriptional regulator previously implicated in MS. ChIP-seq analysis revealed that EBNA2 binding with its interacting human partners RBPJ, EBF1, and PU.1 is highly enriched at MS genetic risk loci, with extensive EBNA2 allelic binding and increased enrichment at MS genetic risk loci in MS-derived cells. Our findings demonstrate that enhanced EBNA2 activity in MS alters human gene expression, chromatin accessibility, and transcription factor binding in an MS-dependent manner. Collectively, this study provides new insights into the molecular mechanisms through which EBV, particularly EBNA2, interacts with host genetic risk to contribute to MS pathogenesis.

## Introduction

Multiple sclerosis (MS) is an immune-mediated demyelinating disease of the central nervous system, with approximately 2.8 million individuals affected globally (Walton et al. 2020). It is the most common cause of non-traumatic neurological disability in young adults (Filippi et al. 2018). MS is caused by a complex combination of genetic and environmental risk factor interactions (Olsson et al. 2017). Genetic factors account for approximately 30% of disease risk, with the unaccounted risk being attributed to environmental factors, including Epstein-Barr virus (EBV) infection (International Multiple Sclerosis Genetics 2019).

Genome-wide association studies have nominated 233 independent risk loci for MS (International Multiple Sclerosis Genetics 2019). HLA-DRB-15*01 is the only MS risk locus with a moderate effect size (odds ratio ∼ 3), with all other variants having small effect sizes (Hollenbach and Oksenberg 2015). Genetic risk variants are strongly enriched within non-coding regions of the genome, including enhancers and promoters (Maurano et al. 2012), suggesting that transcriptional dysregulation is likely key to MS etiology.

The most well-supported environmental factor in the etiology of MS is EBV, a common gammaherpesvirus that infects over 90% of the world’s population. However, in patients with MS, the infection rate is consistently higher (over 99%) (DeLorenze et al. 2006). In 2022, a longitudinal study solidified this relationship by demonstrating that EBV infection is required for MS pathogenesis. In this retrospective study, EBV infection in early adulthood resulted in a 32-fold increased risk of MS, and serum levels of neurofilament light chain, a biomarker of neuronal damage, increased only after EBV infection (Bjornevik et al. 2022).

Most cell types driving MS pathophysiology are immune-related (e.g., B and T cells), and the success of anti-CD20 therapies in reducing both the severity of symptoms and relapse rate in MS suggests that B cells likely play important roles (Hauser et al. 2017). EBV infects both CD21+ B cells and epithelial cells, and EBV-encoded nuclear antigens and membrane proteins can evade the host’s immune system by controlling the host’s cell cycle and preventing apoptosis. EBV can exploit normal B cell differentiation pathways and cause infected B cells to transition from activated B cell blasts to latently infected resting memory B cells that escape the host immune response (Zhao et al. 2011; Thorley-Lawson 2015). Latency III is the most prevalent viral phase during chronic disease, with the EBV transcriptome facilitating immune evasion (Ressing et al. 2015).

Epstein-Barr virus nuclear antigens expressed during latency, particularly EBNA1 and EBNA2, are hypothesized to play substantial roles in multiple sclerosis (MS). EBNA1 shows molecular mimicry with several human proteins, including GlialCAM, which is found in the central nervous system (Lanz et al. 2022). EBNA2 is a viral nuclear protein that serves as a transcriptional coactivator (Cable et al. 2025). It is thought to contribute to MS pathogenesis through its myriad interactions with the human genome, illustrated by highly enriched binding activity and EBNA2-dependent chromatin alterations at many autoimmune disease risk loci (Cable et al. 2025). EBNA2 binds indirectly to human DNA via human-encoded transcription factors (TFs) such as RBPJ, EBF1, and PU.1 (Waltzer et al. 1994; Johannsen et al. 1995; Murata et al. 2016). Our previous work has demonstrated that 44 MS-associated genetic loci contain a genetic variant located within an EBNA2-occupied region of the genome (Harley et al. 2018). EBNA2 extensively rewires the human genome, altering the expression of over 400 human genes and modifying chromatin accessibility at more than 2,000 genomic regions. Importantly, these regions of the human genome with EBNA2-dependent chromatin accessibility and chromatin looping are highly enriched at MS genetic risk loci (Hong et al. 2021).

In this study, we sought to understand how EBV infection leads to altered gene expression in B cells of patients with MS. To this end, we obtained primary B cells from patients with MS along with matched healthy controls (HC). After measuring gene expression and chromatin accessibility in primary B cells, we uniformly infected cells with the B95-8 strain of EBV. In the resulting transformed B cell lines, we measured gene expression, chromatin accessibility, and genomic binding of EBNA2 and its human-encoded TF interaction partners RBPJ, EBF1, and PU.1. Our results establish that EBV infection causes extensive, robust MS-dependent changes to human gene expression and chromatin accessibility. EBNA2 expression levels account for virtually all of these differences, and EBNA2 binds along with its human TF partners at differentially expressed genes and differentially accessible chromatin. Building upon past studies, we demonstrate highly enriched EBNA2 occupancy at MS genetic risk loci in MS patient-derived cells, with increased enrichment in patients with MS relative to healthy controls. Altogether, this study establishes molecular mechanisms across the human genome through which EBV rewires B cells in an MS-dependent manner through EBNA2.

## Results

We recruited individuals representing the largest demographic of patients in the United States: MS women of European ancestry between the age of 25-74 (Filippi et al. 2018). Twelve individuals with clinician-diagnosed MS were recruited along with eleven age-matched healthy controls. The demographics and clinical phenotypes of each individual are provided in Supplemental Table S1. Peripheral blood mononuclear cells (PBMCs) were obtained from each subject. CD19+ primary B cells were isolated and EBV-transformed B cell lines were uniformly generated using standard protocols (see Methods, Supplemental Fig. S1).

### Extensive MS-dependent gene expression differences in B cells after EBV transformation, which can be accounted for by *EBNA2* expression levels

We performed bulk RNA-seq experiments in primary B cells and EBV-transformed B cells from patients with MS and matched controls (see Methods, Supplemental Tables S2 and S3). We observed very few transcriptional differences in the primary B cells of individuals with MS and matched controls (19 total genes) (Fig. 1A). However, in EBV-transformed B cells, we observed 1,060 differentially expressed genes between MS and HC (Fig. 1B). Based on the established role of EBNA2 in the genetic etiology of MS (Mechelli et al. 2015; Ricigliano et al. 2015; Harley et al. 2018; Afrasiabi et al. 2019; Afrasiabi et al. 2020; Keane et al. 2021), we tested the hypothesis that *EBNA2* expression levels, a proxy for EBNA2 activity, might account for these differences. Indeed, *EBNA2* expression levels across cell lines accounted for virtually all MS-dependent differential expression (Fig. 1C). Despite other EBV genes also being dysregulated in MS, no other EBV gene (e.g., *EBNA1*, *EBNA-LP*) could account for as much differential expression as *EBNA2* (Supplemental Fig. S2). Among the altered human genes, EBV-induced molecule 2 (also known as *GPR183*) is a chemotactic receptor that regulates B cell migration (Gatto and Brink 2013). *GPR183* is upregulated in MS EBV-transformed B cells, and *GPR183* expression levels can be explained by *EBNA2* expression levels (Supplemental Fig. S3). *ACKR3,* another gene that is important for B cell chemotaxis, is the most significantly upregulated gene in MS EBV-transformed B cells, and this can also be accounted for by *EBNA2* expression levels (Supplemental Fig. S3). CD40 is a central immune costimulatory receptor that promotes antigen presentation and lymphocyte activation, is implicated in MS neuroinflammation, and is currently being targeted therapeutically to reduce disease activity (Vermersch et al. 2024). While there are no MS-dependent *CD40* expression differences in primary B cells, after EBV transformation *CD40* expression is increased in cells from people with MS relative to controls, with *CD40* expression in the EBV-transformed B cells again being accounted for by *EBNA2* expression levels (Supplemental Fig. S3).

**Figure 1.**
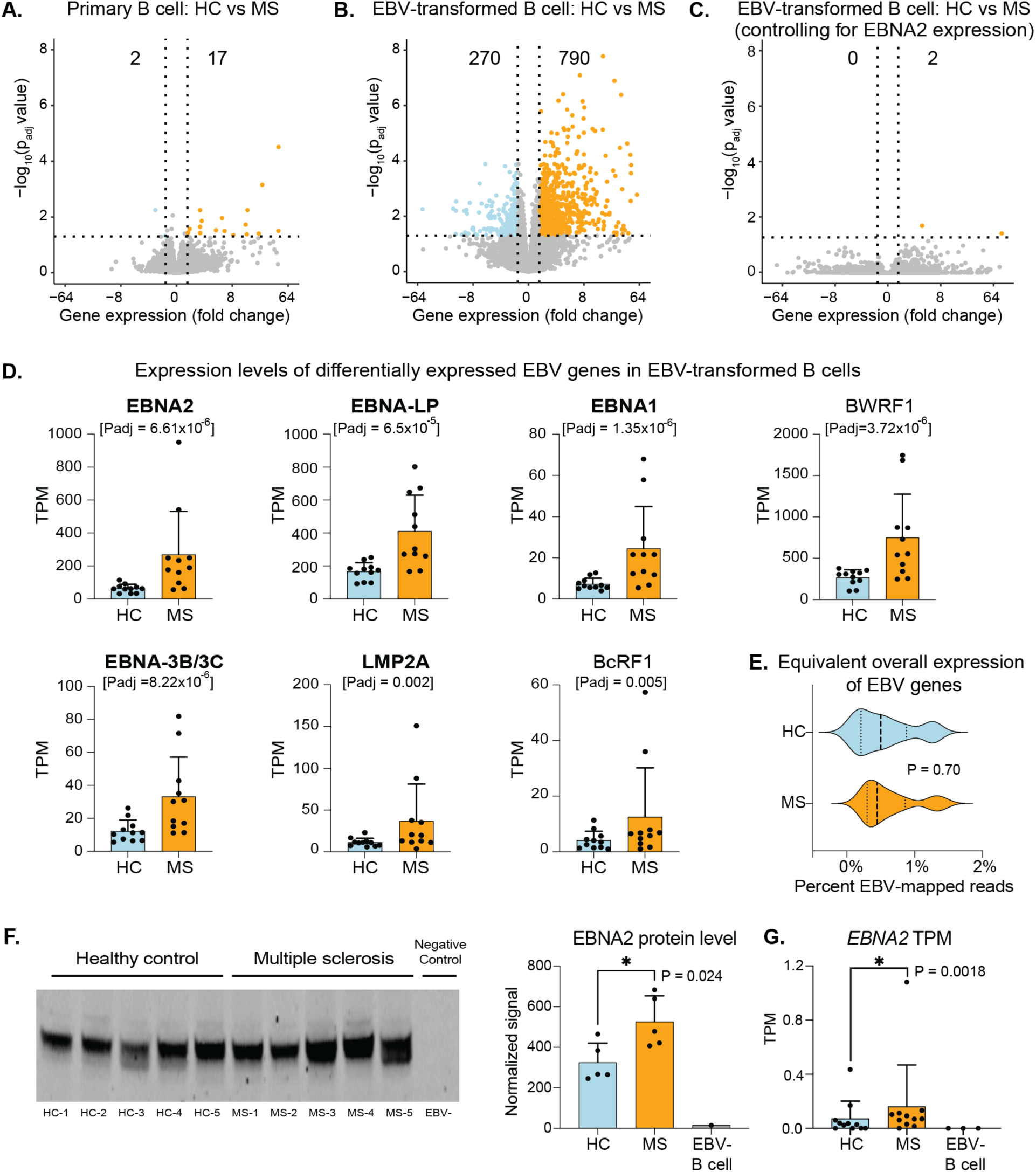
*EBNA2* expression accounts for extensive MS-dependent gene expression differences in B cells after EBV transformation. A-C. Comparison of human gene expression levels (RNA-seq) in healthy controls (HC) and patients with multiple sclerosis (MS). Horizontal dashed lines indicate an adjusted p-value significance threshold of 0.05. Vertical dashed lines indicate a fold-change threshold of 1.5. Results are shown for primary B cells (A), EBV-transformed B cells (B), and EBV-transformed B cells after statistically adjusting for *EBNA2* expression levels (C). D. All significant differentially expressed EBV genes in EBV-transformed B cells from HC and patients with MS. Gene expression (RNA-seq) is shown in transcripts per million (TPM). Each dot represents a cell line derived from one individual. Error bars indicate standard deviation. EBV genes in bold are expressed during latency III. E. The total percent of RNA sequencing reads mapped to the EBV genome in EBV-transformed B cells derived from HC and MS. Vertical dashed lines denote quartiles. F. Western blot for EBNA2 protein in HC and MS cell lines (left) and quantification of EBNA2 protein (right). For full blots of EBNA2 and total protein stains, see Supplemental Fig. S4. G. *EBNA2* gene expression levels (RNA-seq) in primary B cells in transcripts per million (TPM). Each dot represents primary B cells isolated from one individual. A B cell line without EBV was used as a negative control. Error bars indicate standard deviation.

Consistent with these results, *EBNA2* is the second most significant differentially expressed gene of the 84 EBV-encoded genes, with significantly higher levels in people with MS (p adj = 6.61E-06) (Fig. 1D). Among the seven differentially expressed EBV genes, five are expressed during latency III, including *EBNA2*. We confirmed that total EBV gene expression levels are not driving these differences (Fig. 1E). We next performed western blots for EBNA2 in 5 HC and 5 MS cell lines and found significantly increased levels of EBNA2 protein in MS cells (Fig. 1F, Supplemental Fig. S4). Taken together, we conclude that *EBNA2* expression levels can account for the vast majority of MS-dependent gene expression differences in EBV-transformed B cells, independent of overall EBV gene expression levels.

To rule out the possibility that differential prior exposure to EBV might play a role in the observed gene expression differences, we examined our cohort for evidence of prior EBV infection through the detection of EBV-derived RNA or DNA, and through serological evidence of immune responses to EBV. As expected, based on worldwide EBV infection rates (∼95%), everyone in our cohort had evidence of prior infection by EBV (Supplemental Table S1, Supplemental Fig. S5).

We next hypothesized that MS-dependent differences in the abundance of individual B cells that were initially transformed by EBV could drive the observed differential expression (Fig. 1B). To test this hypothesis, we performed single cell RNA-sequencing with B cell receptor (BCR) enrichment in cells obtained from 5 patients with multiple sclerosis and 5 healthy controls (Supplemental Fig. S6). To benchmark our single cell dataset against prior studies, we assessed the differential expression of the BCR subunit immunoglobulin heavy chain. Consistent with previous findings in peripheral B cells (von Budingen et al. 2012), *IGHV3-74* is the most upregulated immunoglobulin gene in MS (Supplemental Fig. S6). We then asked if differences in B cell clonality might account for the observed gene expression differences. Because each B cell’s BCR is uniquely rearranged, we used unique BCR sequences as a method to measure the clonality of each cell line derived from HC and MS. Based on these analyses, we did not find evidence of differences in the clonality of cell lines between HC and MS after EBV transformation (Supplemental Fig. S6).

Next, we tested whether genetic risk might account for some of the observed gene expression differences. To this end, we performed whole genome sequencing to enable imputation of the strongest genetic risk locus (HLA-DRB1*15:01) and calculation of polygenic risk scores (PRS). The distribution of HLA-DRB1*15:01 alleles was virtually identical across HC and MS in our cohort (Supplemental Fig. S7), which is consistent with the effect size and the statistical power requirements to identify the original genetic association (Schmidt et al. 2007). We next calculated MS PRS for each member of our cohort to measure the cumulative genetic burden of risk in each person. As expected, PRS was higher on average in the individuals with MS compared to HC (Supplemental Fig. S7). Furthermore, PRS accounted for 16.4% of differential human gene expression, emphasizing the role of genetics and environment in the observed differences of MS-derived EBV-transformed B cells compared to controls (Fig. 1).

We next sought to further examine the small number of differences in primary B cells between patients with MS and controls. These differences could explain the subsequent large number of transcriptional differences after transformation with EBV (Fig. 1B). EBV can only transform B cells that express CD21. We therefore examined a published single cell RNA-seq dataset from an independent cohort of 3 HCs and 10 people with MS (Ramesh et al. 2020) and found that transitional and naïve B cells express the highest amount of *CD21* among B cell subsets, as expected (Supplemental Fig. S8). We found no differences in the proportion of B cells within the CD21+ transitional and naïve B cell compartments between HC and MS (Supplemental Fig. S8). We identified ten genes that are differentially expressed in MS relative to controls within the CD21+ transitional/naïve B cell compartment, including *BACH2, AFF3,* and *BANK1*, which have all previously been implicated in MS (Supplemental Fig. S8; Supplemental Table S4) (International Multiple Sclerosis Genetics et al. 2011; Itoh-Nakadai et al. 2014; Bae and Lee 2017; Tsukumo et al. 2022). *FANCC* is upregulated in MS CD19+ primary B cells (Supplemental Table S3). A CRISPR screen demonstrated that *FANCC* expression is essential for EBNA2-dependent transformation of primary B cells to EBV-transformed cell lines (Ma et al. 2017). This gene also demonstrated increased expression in CD21+ transitional/naïve B cells in the independent set of HC and patients with MS for which we have single cell RNA-seq data, which is trending toward significance (5.2-fold increase, p-value = 0.17). Taken together, these data support a model in which primary B cells from patients with MS have higher expression of *FANCC*, a gene critical for EBV-dependent immortalization of B cells, along with several other genes with prior links to MS and B cell biology. Strikingly, while the primary B cells from patients with MS and controls had equivalent levels of RNA mapped to the EBV genome (Supplemental Fig. S5), primary B cells from patients with MS had significantly higher expression of EBNA2 compared to controls (Mann-Whitney U test, p = 0.0018) (Fig. 1G). The identification of enhanced EBNA2 expression in the primary B cells before transformation for this study suggests a fundamental difference in the B cell compartment of patients with MS relative to controls, a finding consistent with prior studies (Ramesh et al. 2020; Soldan et al. 2024; Sarkkinen et al. 2025).

### EBV-transformed B cells from patients with MS have distinct chromatin accessibility patterns compared to controls

Gene expression differences are often driven by differences in chromatin accessibility. To identify chromatin accessibility differences between people with MS and healthy controls, we performed ATAC-seq in primary and EBV-transformed B cells in six HC and six individuals with MS. The resulting data met our stringent quality control criteria, which are based on the ENCODE consortium standards (Luo et al. 2020; Hitz et al. 2023) (Supplemental Table S2). Consistent with the gene expression analysis, we identified only minimal differences in chromatin accessibility in the primary B cells of HC compared to MS (Fig. 2A, Supplemental Table S5). Also consistent with the gene expression analysis, transformation with EBV resulted in large differences between MS and HC, with 1,197 peaks that were stronger in MS and 287 that were stronger in HC (Fig. 2B, Supplemental Table S5). Accounting for *EBNA2* expression levels again removed the vast majority of these disease-dependent differences (Fig. 2C).

**Figure 2.**
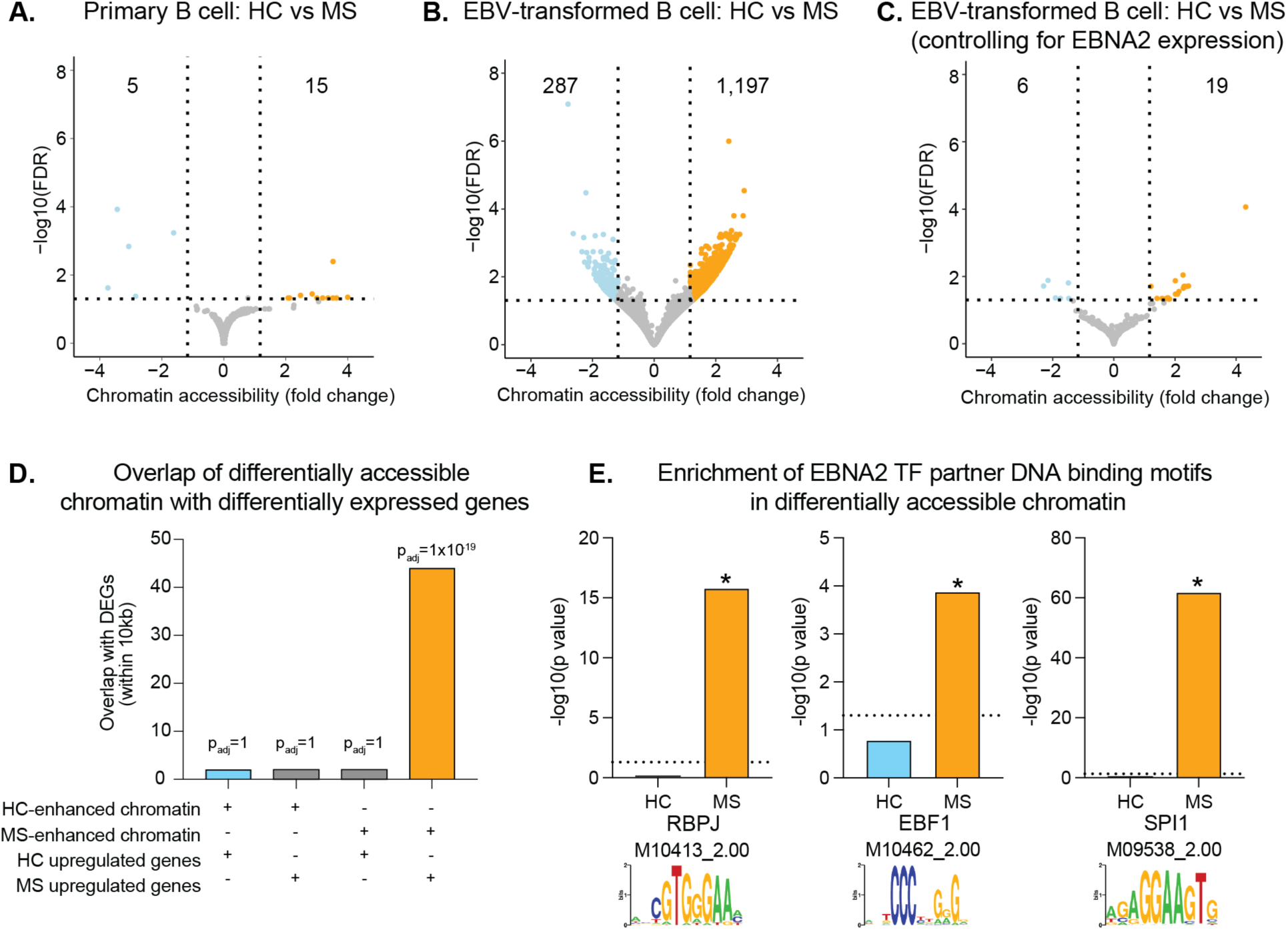
*EBNA2* expression accounts for MS-dependent chromatin accessibility differences in EBV-transformed B cells. A-C. Comparison of chromatin accessibility (ATAC-seq) in healthy controls (HC) and patients with multiple sclerosis (MS). Horizontal dashed lines indicate an adjusted p-value significance threshold of 0.05. Vertical dashed lines indicate a fold-change threshold of 1.5. Results are shown for primary B cells (A), EBV- transformed B cells (B), and EBV- transformed B cells after statistically adjusting for *EBNA2* expression levels (C). D. Overlap of all possible pairs of differentially accessible chromatin (ATAC-seq) and differentially expressed genes with 10 kb padding on either side of the TSS (RNA-seq). Each set of chromatin and differential genes were either up in HC or MS. The statistical enrichment of the overlap (measured by RELI, see Methods) is provided as an adjusted p-value on top of each bar. E. DNA binding motif enrichment of exemplary motifs for EBNA2 TF partners RBPJ, EBF1, and SPI1 (PU.1) in accessible chromatin. Enrichment is calculated as % foreground motifs divided by % background motifs (i.e., the percent of randomly selected genome sequences, matching GC content). Asterisks indicate significant motif enrichment (P < 0.05), as calculated by HOMER.

We next examined the concordance between chromatin accessibility changes and gene expression changes. To this end, we calculated the statistical significance of the overlap between differentially expressed genes and differentially accessible chromatin using our RELI algorithm (Harley et al. 2018) (see Methods, Supplemental Table S6). Importantly, stronger chromatin accessibility in EBV-transformed B cells from patients with MS corresponded with higher gene expression in these cells (Fig. 2D, right bar). This relationship was not observed in any of the other possible pairwise relationships; for example, we did not observe correspondence between closed chromatin and activated gene expression (Fig. 2D, other bars). As EBNA2 does not directly bind DNA, we also calculated the enrichment of the DNA binding motifs of its TF interaction partners RBPJ, EBF1, and PU.1 (encoded by *SPI1*) in differentially accessible chromatin. The motifs for all three TFs were significantly enriched in chromatin with enhanced accessibility in MS, but not in chromatin that is more accessible in HC (Fig. 2E), supporting a model where increased EBNA2 levels result in increased human gene expression levels through increased chromatin accessibility at regions bound by EBNA2 with its human TF interaction partners.

### Binding of EBNA2 and human TF interaction partners in EBV-transformed B cells

Our data reveal extensive MS-dependent chromatin accessibility and gene expression that can be explained by EBNA2 expression levels. To further understand the role of EBNA2 and its TF interaction partners in these differences, we used ChIP-seq to measure the genome-wide binding of EBNA2, RBPJ, EBF1, and SPI1 (PU.1) in five HC and five MS EBV-transformed B cell lines obtained from cohort members with corresponding gene expression and chromatin accessibility measurements. The resulting data met our stringent quality criteria that were derived from the ENCODE consortium (Luo et al. 2020; Hitz et al. 2023) (Supplemental Table S2).

We observed thousands of robust ChIP-seq peaks, with an elevated average peak count in MS compared to controls for all four proteins (Fig. 3A). In cells from patients with MS, EBNA2 binding significantly overlapped genes with increased expression in MS-derived EBV transformed B cells (Fig. 3B), binding within 10kb of the TSS for over 25% of genes with MS-dependent expression (Supplemental Table S6). Likewise, the three EBNA2 TF partners also demonstrated significant overlap with genes whose expression increased in MS-derived cell lines (Fig. 3B). When accounting for co-occurrence of EBNA2 and human TF ChIP-seq peaks in cells from the same individual with MS, we again identify the strongest enrichment of overlap of EBNA2-TF pairs at genes with increased expression in MS-derived cells (Fig. 3C).

**Figure 3.**
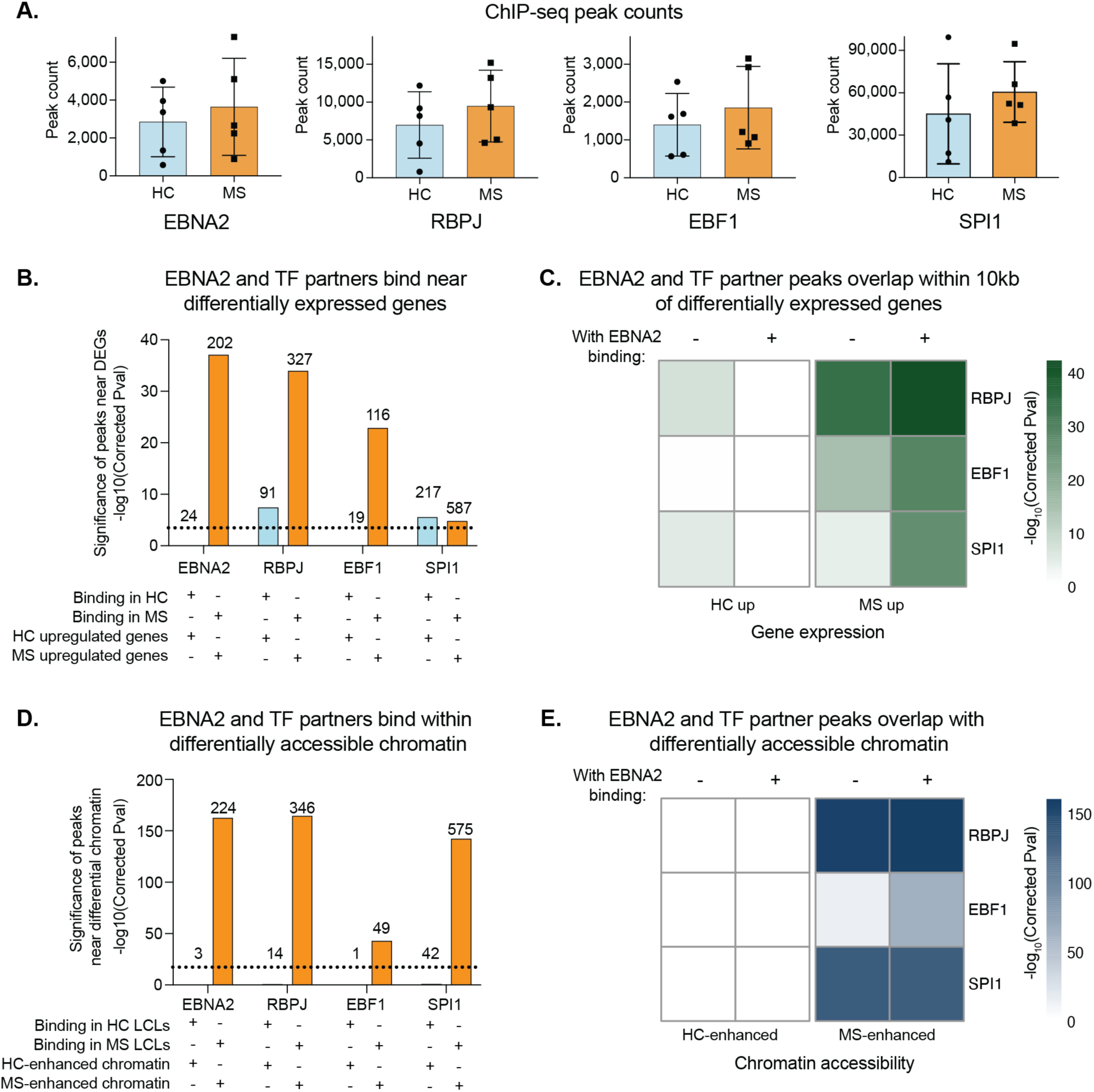
EBNA2 and TF partners bind near genes upregulated in MS EBV-transformed B cells and MS-enriched open chromatin. A. Total number of ChIP-seq peaks for each experiment. Each dot indicates one EBV-transformed cell line from a healthy control (HC) or person with multiple sclerosis (MS). Error bars indicate standard deviation. B. Overlap of all possible pairs of EBNA2, RBPJ, EBF1, or SPI1 (ChIP-seq) and differentially expressed genes with 10 kb padding on either side of the gene (RNA-seq). A combined set of ChIP-seq peaks found in any of the HC or MS-derived EBV-transformed B cells was used for each protein. Differentially expressed genes were higher in either HC or MS-derived EBV-transformed B cells. The statistical enrichment of the overlap (measured by RELI, see Methods) is provided as an adjusted p-value on the y-axis. Dotted line indicates significance threshold. The number of overlaps is provided on top of each bar. C. Co-occurring binding of EBNA2 with each human transcription factor (RBPJ, EBF1, or SPI1) was called within each EBV-transformed B cell line derived from HC and individuals with MS, using ChIP-seq peaks from the same cell line. For each EBNA2–TF pair, we then compiled a single, pooled set of co-occurrence sites by taking the union of co-occurring peaks observed in any HC- or MS-derived cell line. These co-occurrence peak sets were tested for enrichment near differentially expressed genes (genes upregulated in HC or upregulated in MS). The heatmap shows the significance of each enrichment test, quantified by RELI, for each EBNA2–TF pair and gene set comparison. D. Overlap of all possible pairs of EBNA2, RBPJ, EBF1, or SPI1 (ChIP-seq) and differentially accessible chromatin (ATAC-seq). A combined set of ChIP-seq peaks found in any of the HC or MS-derived EBV-transformed B cells was used for each protein. Chromatin was significantly more accessible in either HC or MS-derived EBV-transformed B cells. The statistical enrichment of the overlap (measured by RELI, see Methods) is provided as an adjusted p-value on the y-axis. The number of overlaps is provided on top of each bar. E. Co-occurrence of EBNA2 and either RBPJ, EBF1, or SPI1 was identified in each EBV-transformed B cell line in both HC and MS. A combined set of co-occurring peaks found in any of the HC or MS-derived cell lines was used. Each set of peaks (ATAC) was significantly more accessible in either HC or MS. The heatmap depicts the significance of the overlap (as measured by RELI) for each pairwise comparison.

Next, we compared chromatin accessibility changes with the binding of EBNA2 and identified statistically significant overlap in cells derived from patients with MS and MS enhanced chromatin (Fig. 3D). We also observed strong statistical enrichment for the co-occupancy of EBNA2 with TF partners RBPJ, EBF1, and SPI1 in MS-derived cells within MS-enhanced chromatin, but not for chromatin that is more accessible in HC (Fig. 3E).

In summary, upon assessing EBNA2 binding in cell lines derived from HC, the overlap with HC-dependent genes and HC-enhanced chromatin was minimal, with far less significance relative to MS. These results highlight the fact that many more genes are increased in MS (790 genes) relative to HC (270 genes) (Fig. 1B) and many more ATAC-seq peaks are enhanced in MS (1,197) relative to HC (287) (Fig. 2B). Altogether, these results reveal a model of enhanced EBNA2 activity in MS compared to HC, leading to alterations in EBNA2 binding, human TF binding, human chromatin accessibility, and human gene expression.

### EBNA2 has increased occupancy at MS genetic risk loci

We next asked whether the enhanced EBNA2 activity in MS-derived cells preferentially targets known MS genetic risk loci. We previously discovered that EBNA2 occupies 44 of the 109 independent MS-associated genetic loci tested (Harley et al. 2018). With the number of established independent genetic risk loci more than doubling in the past few years, and the number of available EBNA2 ChIP-seq datasets likewise increasing (Hong et al. 2021; Viel et al. 2024), we next sought to determine updated EBNA2 occupancy of the expanded set of risk loci in MS-derived cells. Using the RELI algorithm (Harley et al. 2018) and data from the present study, we find that EBNA2 occupies 33 and 25 MS risk loci in EBV-transformed B cells from patients with MS and HC, respectively (Fig. 4A, Supplemental Table S6). Accordingly, the enrichment of EBNA2 at MS risk loci is more significant in MS derived cells (adjusted p-value = 5.81E-14) compared to those from HC (adjusted p-value = 2.63E-09). While most loci and risk variants were shared between HC and MS, many had EBNA2 occupancy only in MS-derived (13 loci; 25 risk variants) or HC-derived (5 loci; 9 risk variants) cells (Fig. 4B).

**Figure 4.**
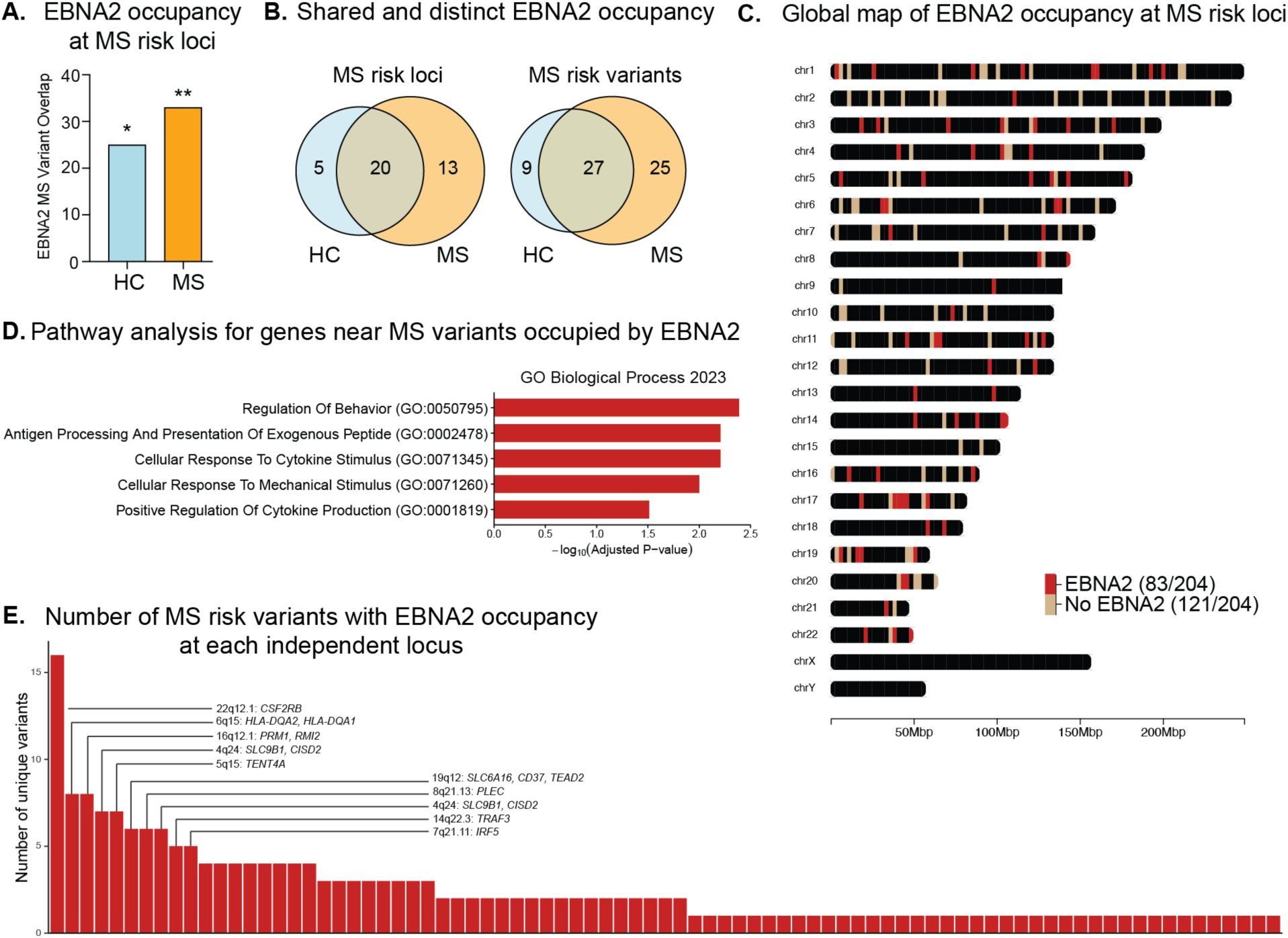
EBNA2 occupancy at MS genetic risk loci. A. EBNA2 occupancy at MS risk loci in HC and MS EBV-transformed B cells. Count of EBNA2 ChIP-seq peaks overlapping one or more MS risk variants within each independent locus from HC (left) or MS (right). Overlap significance (as calculated by RELI, see Methods) is indicated with asterisks (*, p = 2.63E-09; **, p = 5.81E-14). B. Comparison of EBNA2 MS risk locus occupancy in HC- or MS-derived cells. (left) Number of MS genetic risk loci where an EBNA2 ChIP-seq peak overlaps with one or more MS risk variant. (right) Total number of these genetic variants (SNPs) (i.e., the counts include multiple variants within a locus). C. Chromomap depicting EBNA2 occupancy at MS risk loci in EBV-transformed B cells. EBNA2 ChIP-seq peaks were collected from a variety of sources, including this study (see Methods). D. Pathway enrichment (Gene Ontology Biological Process) of genes with TSS within 10kb of MS risk loci bound by EBNA2. E. Number of MS risk variants with EBNA2 occupancy at each independent locus.

To further expand the set of EBNA2 human genomic targets, we performed ChIP-seq in 21 additional cell lines in a wide range of B cell types and contexts (Supplemental Table S2). Combining these data with all publicly available EBNA2 genomic binding data, we created an updated global map of EBNA2 occupancy, which reveals EBNA2 occupancy at 83 of the 204 known MS risk loci (Fig. 4C). Pathway enrichment analysis reveals that EBNA2 binds at MS risk loci near genes related to pathways involved in key B cell functions such as antigen presentation, response to cytokine, and cytokine production (Fig. 4D, Supplemental Table S7). While most EBNA2-bound MS risk loci had a single EBNA2 binding event overlapping an MS variant, a subset of MS risk loci had multiple variants bound by EBNA2, such as the *CSF2RB* locus (n=16 EBNA2-bound variants) and *HLA* and others, implicating EBNA2 binding at 204 unique MS variants within 83 independent MS genetic risk loci (Supplemental Table S6) [21].

### Genotype-dependent EBNA2 occupancy at MS risk variants

Our analyses indicate that EBNA2 occupancy is significantly enriched at MS risk variants. We next wanted to investigate allele-dependent EBNA2 binding events at these loci. To this end, we used our MARIO pipeline to systematically identify MS risk allele-dependent binding of EBNA2 (Harley et al. 2018). In brief, MARIO identifies heterozygous genetic variants within ChIP-seq peaks with significantly different read counts between the two alleles (see Methods). We combined the EBNA2 ChIP-seq experiments produced in this study with publicly available data and identified allele-dependent EBNA2 binding events at MS risk variants. In total, we identified 38 instances of allele-dependent EBNA2 binding to 18 unique MS risk variants in EBV-transformed B cells (Fig. 5A, Supplemental Table S8). Among these nominated risk variants, rs17035378 and rs8072391, which both have stronger EBNA2 binding to the MS risk allele, are eQTLs in EBV-transformed B cells for *PLEK* and *STAT3*, respectively (Fig. 5B). When all MS risk variants that are bound by EBNA2 in a genotype-dependent manner are connected through eQTL analysis, including those from this study and previously published variants, a network of genes emerges containing a significant number of interactions (STRING-db analysis, p = 8.73 e-5, Fig. 5C). From the interaction network, we identify a set of genes involved in innate immune response and a second set with roles in cytokine signaling (Fig. 5C). Together, these analyses reveal a large core set of human genes whose expression is likely altered by allele-dependent EBNA2 binding to MS-associated genetic risk variants, revealing important gene/environment disease mechanisms for potential therapeutic targeting.

**Figure 5.**
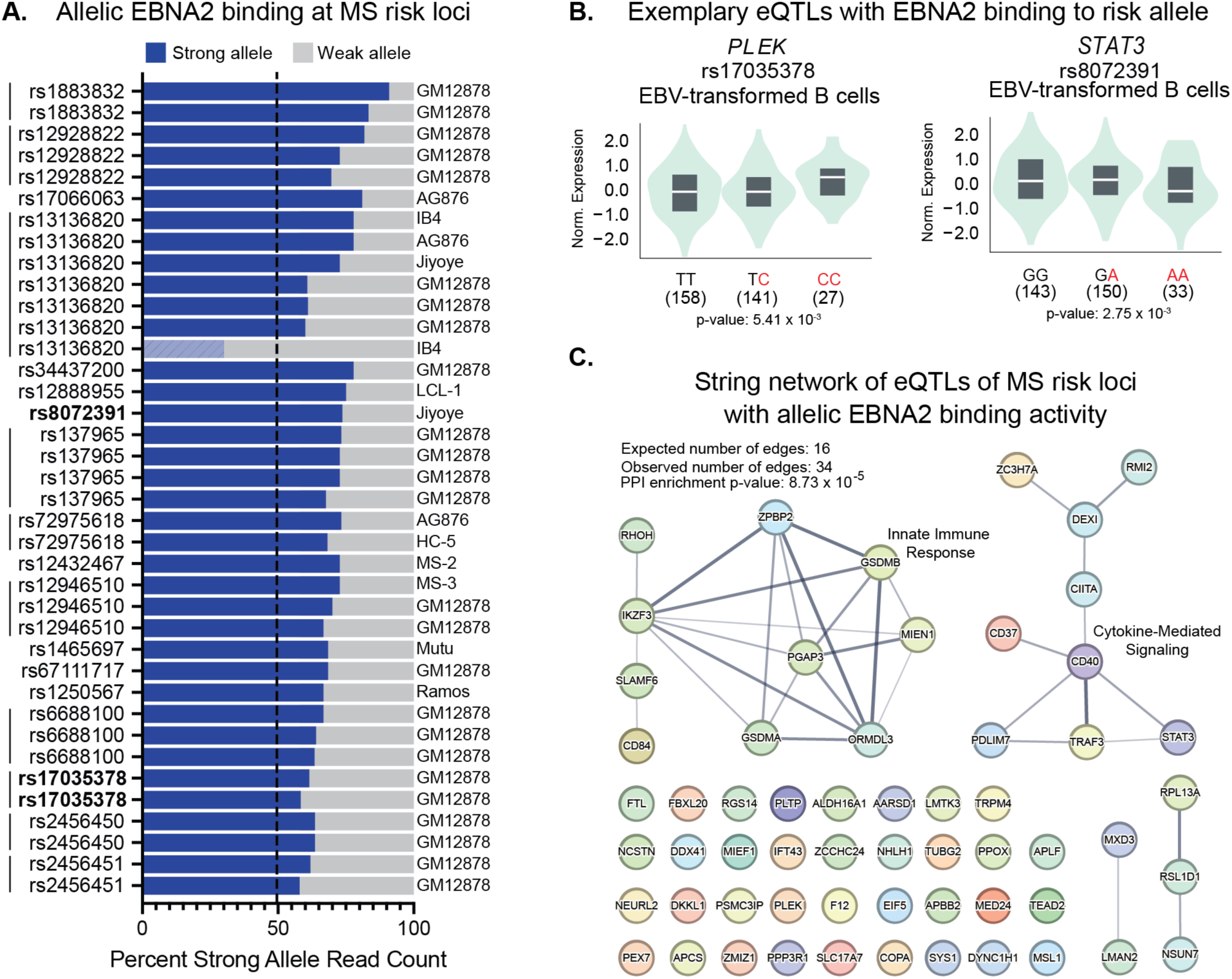
Allelic EBNA2 occupancy at MS risk variants. A. MS-associated genetic risk variants with allele-dependent EBNA2 binding. Each variant is heterozygous and located within an EBNA2 ChIP-seq peak to identify allele-dependent occupancy. The x-axis indicates the percent of reads that map to the strong allele vs the weak allele. A strong allele is defined as the allele preferred in the majority of experiments for that variant. A single variant had discordant allelic results in one out of seven experiments (indicated in hatches). Variants shown in eQTL plots in panel B are bolded. Identical variants in different datasets are connected with vertical bars. B. Genotype-dependent expression of *STAT3* and *PLEK* at MS risk variants with allelic EBNA2 binding (identified in GTeX). MS risk alleles are indicated in red. C. eQTLs at MS variants with allelic EBNA2 binding were identified in the eQTL catalogue. A network of these eQTLs was produced using STRING.

## Discussion

In this study, we show that the EBV-encoded transcriptional regulator EBNA2 has increased activity in EBV-transformed B cells derived from patients with MS compared to controls, and that this heightened activity amplifies MS-relevant transcriptional and chromatin programs. We observe extensive MS-dependent changes in gene expression and chromatin accessibility that are largely explained by higher EBNA2 expression in MS-derived EBV-transformed B cells (Fig. 1). To explain this phenomenon, we systematically evaluated potential confounders, including B cell clonality, prior EBV exposure, the proportion of CD21+ B cells, overall EBV gene expression, cumulative genetic burden, and HLA-DRB1*15:01 status; none accounted for the MS-dependent gene expression as effectively as *EBNA2* expression levels. Generally, increased EBNA2 expression levels tend to lead to increases in human gene expression levels, affecting genes involved in MS-relevant pathways such as *GPR183* (B cell migration and crosstalk with T cells), *ACKR3* (B cell adhesion and migration), and *CD40* (B cell proliferation and survival). Our MS-derived cells also reveal differential binding of EBNA2, differential chromatin accessibility, and genotype-dependent binding of EBNA2 at MS risk loci.

Overall, we observed dramatic differences between primary B cells before and after EBV-transformation. Prior to EBV infection, we observed only 19 human genes with altered expression between MS and controls, with many involved in B cell activation such as *FANCC*. In an independent single-cell dataset, *FANCC* was again higher in CD21+ transitional/naïve B cells from patients with MS, and a prior CRISPR screen identified *FANCC* as essential for EBV-mediated B cell transformation (Ma et al. 2017). Together, these findings suggest that subtle pre-existing differences in activation programs of CD21+ B cells may prime MS-derived B cells for enhanced EBV transformation and EBNA2-dependent reprogramming.

Subsequent to EBV transformation, we observed substantial differences between patients with MS and matched controls on multiple levels: gene expression levels, chromatin accessibility, and human TF binding. EBNA2 is the most significant differentially expressed EBV gene in MS-derived cells (Fig. 1D), EBNA2 protein levels are higher in MS-derived cells (Fig. 1F), and EBNA2 expression level can account for the extensive gene expression differences seen in HC and MS cell lines (Fig 1C.). Occupancy of EBNA2 and its human TF binding partners is highly enriched for human genes with increased chromatin accessibility and increased expression levels in patients with MS (Fig. 3B, 3D). Many of these EBNA2 binding events also coincide with MS risk loci (Fig. 4), consistent with prior studies from several groups including our own (Ricigliano et al. 2015; Harley et al. 2018; Hong et al. 2021; Viel et al. 2024). In total, EBNA2 occupies 33 and 25 risk loci in EBV-transformed B cells from patients with MS and HC, respectively (Fig. 4A). While the majority of loci were shared between HC and MS, many loci had EBNA2 occupancy only in MS- (13 loci) or HC-derived (5 loci) cells (Fig. 4B). We also identify allelic EBNA2 occupancy at 18 MS risk variants, strengthening our current understanding of MS genotype-dependent biolog. Compared to prior EBNA2 studies that relied on established cell lines, our use of patient-derived EBV-transformed B cells, coupled with allele-dependent analyses, provides direct evidence that EBNA2 engages MS risk loci in a disease- and genotype-dependent manner.

Our study uses the B95-8 laboratory strain of EBV to infect primary B cells derived from controls and patients with MS, providing a controlled model of latent EBV infection in human B cells with known genetic background and disease status. Consistent with this model, a recent study in systemic lupus erythematosus demonstrated that EBV preferentially infects autoreactive antinuclear B cells and reprograms them into antigen-presenting cells that activate T peripheral helper cells and propagate pathogenic T and B cell responses (Younis et al. 2025). These data provide direct evidence that rare EBV infection events in autoreactive B cells can initiate systemic autoimmunity, aligning with our proposed model for EBV–B cell interactions in MS. They also complement a recent study that used spontaneous lymphoblastoid cell lines to interrogate endogenous EBV in MS-derived B cells, revealing EBV lytic gene expression in active disease compared to both controls and patients with stable disease (Soldan et al. 2024). Those data, together with prior serologic evidence of EBV reactivation during active MS (Wandinger et al. 2000; Farrell et al. 2009), highlight dysregulated lytic infection. In contrast, our work focuses on latent EBV infection under controlled viral and disease conditions, showing that even in the absence of overt lytic reactivation, heightened EBNA2 activity can remodel chromatin and gene expression at MS risk loci. Together, these studies support a model in which impaired control of both latent and lytic EBV infection in B cells contributes to MS pathogenesis. Importantly, both studies reveal reduced control of EBV B cell infection as a potential driver of multiple sclerosis.

The goals of this study are to clarify how interactions between genetic variants and EBNA2 influence MS disease processes, and to define the pathways whose regulation and cell-fate decisions differ in people with MS compared to those without MS. Even common viruses like EBV can contribute to rare non-communicable diseases such as MS. This is likely driven by a combination of host genetics, stochastic events, and environmental factors. In the case of EBV, only a modest minority of people who are infected experience symptoms and develop mononucleosis (Crawford et al. 2006). Likewise, of the ∼95% of people with prior EBV infection, the vast majority will not develop subsequent autoimmune diseases (Ascherio and Munger 2015). While up to 50% of this variation may be due to genetics (International Multiple Sclerosis Genetics 2019), stochasticity also likely plays a role in determining when EBV will lead to MS or other autoimmune diseases. Because only a tiny fraction of B cells are infected with the virus, and the likelihood of those infected B cells possessing a disease-driving BCR is remote (SoRelle et al. 2022), it is possible that the disease-inducing BCR and EBV never occur within the same cell to initiate the cascade of EBNA2-induced changes we found. Likewise, environmental factors like vitamin D level and timing of initial EBV infection will also impact the likelihood of progressing to an MS disease state. Additional studies such as this one are needed to explore these triggers further, for EBV and other viruses.

Our findings reveal a key role for the EBV-encoded EBNA2 protein in transcriptional regulation of MS-derived B cells. Heightened EBNA2 expression in MS-derived cells compared to healthy controls is accompanied by disease-dependent changes in EBNA2 binding, gene expression, and chromatin accessibility. In total, we observe EBNA2 binding at 204 MS genetic risk variants, encompassing 83 distinct MS genetic risk loci, including allele-dependent EBNA2 binding events to 18 different MS-associated variants. Collectively, these results reveal an extensive network of EBNA2-induced MS risk-dependent gene regulatory alterations, providing robust mechanistic targets for therapeutic intervention.

## Methods

### Ethics statement

This study was approved by the Cincinnati Children’s Hospital Institutional Review Board (IRB #2017-0430) in March of 2017. Both participants with Multiple Sclerosis (MS) and Healthy Controls (HC) signed informed consent forms prior to participation.

### Patient recruitment

Patients with a diagnosis of MS (n=12) were recruited from the Waddell Center for Multiple Sclerosis at the University of Cincinnati or through a study advertisement sent out to Cincinnati Children’s Hospital employees. To reduce heterogeneity, the study inclusion criteria for subjects with MS was restricted to European American women between the ages of 25 and 74, which is representative of the largest MS demographic. Eleven age- and ancestry- matched controls were recruited from a study advertisement sent out to Cincinnati Children’s Hospital employees. Supplemental Table S1 provides the age, sex, self-reported race, ethnicity, and prior EBV infection status of each subject in this study.

### Primary B cell isolation

Immediately after sample collection, peripheral blood mononuclear cells (PBMCs) were isolated from blood samples by Ficoll density gradient centrifugation (mean of 132.3 million +/- 84.9 million cells). CD19+ primary B cells were isolated from PBMCs using a Miltenyi negative selection B cell isolation kit (Miltenyi Biotec 130-091-151).

### ELISA assays

ELISA assays were performed to evaluate prior history of EBV infection. ELISAs for anti-EBNA1 IgG (cat #EI 2793-9601 G) and EBV Capsid Antigen IgG (cat #EI 2791-9601 G) were performed according to the manufacturer’s protocol.

### EBV infection of primary B cells

B95-8 EBV was prepared from cell supernatants after culture in 10% FBS-supplemented RPMI-1640 medium for 2 weeks. Viral suspension was filtered twice with 0.45 µm Millipore filters. The concentrated viral stocks were stored at −80°C. Primary B cells were infected with 1 mL viral stock based on infection optimization assays and incubated for 3 h for virus adsorption. After infection, cells were washed and cultured. Within the first month, we confirmed EBV infection by morphological changes and blasting of the cells. Cells were collected for experiments after at least five passages.

### RNA-seq

Total RNA was extracted using the mirVANA Isolation Kit (Ambion) from 11 primary B cells and 11 EBV-transformed B cell lines and DNase treated with the NucleoSpin® RNA Clean-up XS Kit (MACHEREY-NAGEL). RNA libraries were prepared using the SMARTer® Stranded Total RNA-Seq Kit v2 (Takara Bio USA, Inc.), following the manufacturer’s protocol, and sequenced 2x150 bp on an Illumina NovaSeq 6000 at Cincinnati Children’s Hospital Medical Center (CCHMC) Genomics Sequencing Facility (RRID:SCR_022630).

RNA-seq data were analyzed using the nf-core/rnaseq pipeline (version 3.11.1) (doi: 10.1038/s41587-020-0439-x). Adapter sequence contents were removed using cutadapt (Martin 2011) (trimgalore version: 0.6.7) (https://journal.embnet.org/index.php/embnetjournal/article/view/200). RNA-seq reads were aligned to the GRCh38 genome augmented with EBV B95-8 coding sequences (ASM240226v1, GCF_002402265.1) using Spliced Transcripts Alignment to a Reference (STAR, version: 2.7.9a) (https://doi.org/10.1093/bioinformatics/bts635). Ribosomal RNA was removed using the Silva Library (https://www.arb-silva.de/) and SortmeRNA v 4.3.4 (https://bioinfo.lifl.fr/RNA/sortmerna/). All RNA-seq datasets used in this study passed our quality control checks, which were facilitated by FastQC (version: 0.11.9) (http://www.bioinformatics.babraham.ac.uk/projects/fastqc). Specifically, RseqQC (Wang et al. 2012), Qualimap (Garcia-Alcalde et al. 2012; Okonechnikov et al. 2016), dupRadar (Sayols et al. 2016), and Preseq (https://github.com/smithlabcode/preseq) were used to assess the quality of the datasets. GENCODE (v. 40) (Frankish et al. 2019) annotations for GRCh38 with EBV coding sequence annotations were used to calculate transcript abundance estimates with Salmon (v.1.10.1) (https://doi.org/10.1038/nmeth.4197). The tximport package (https://doi.org/10.12688/f1000research.7563.2) was used to pre-process quantifications and DESeq2 (Love et al. 2014) was used to perform differential gene expression analysis. Differentially expressed genes were defined as those genes with a fold-change difference greater than 1.5 and an FDR of less than 0.05. DESeq2 output was filtered to only include genes with transcript per million (TPM) values greater than one in at least three replicates of MS or HC cell lines. Variables accounting for gene expression levels (e.g., the expression level of the EBNA2 gene) were statistically controlled for using the DESeq2 design formula “ ∼log(gene TPM) + disease_status”.

### Whole genome sequencing and variant calling

Whole genome sequencing was performed on all MS- and HC-derived cells used in this study. DNA was isolated using a PureLink Genomic DNA Kit (Thermo Fisher #K182001). Libraries were sequenced on an Illumina NovaSeq to generate 100-base paired-end reads. Whole genome sequenced files were mapped to the hg38 genome and variants were called using the Sarek v3.4.1 pipeline (Garcia et al. 2020; Hanssen et al. 2024). The Sarek pipeline offers multiple programs for alignment, variant calling, and annotation. These samples were aligned with BWA-mem, variants were called with GATK HaplotypeCaller, and annotated with VEP. In addition, joint variant calling was enabled with the “joint_germline” parameter. To determine prior exposure to EBV, the fraction of reads aligning to the EBV genome was assessed by counting the total number of mapped reads in the duplicate marked CRAM file and comparing to the number of reads mapped to the “chrEBV” contig included as a decoy sequence in the hg38 genome.

### Western Blot

Cells were lysed and sonicated, and protein concentration was determined using a BCA protein assay. 10 μg of lysates were loaded in 4-12% NuPAGE Bis-Tris gels, run in MOPS buffer, and transferred to a nitrocellulose membrane using the iBlot machine (Thermo Fisher Scientific). The membranes were blocked for 1 hour in Intercept Blocking Buffer at room temperature and incubated overnight at 4°C in Intercept Blocking Buffer + 0.1% Tween 20 with anti-EBNA2 (MABE8, Sigma-Aldrich) antibody diluted to 1:1000. The next day, the membranes were washed three times for 5 minutes with PBS + 0.1% Tween 20. The washed membranes were then incubated with rotation for 45 minutes with fluorescent secondary antibodies (diluted to 1:20,000 in Intercept Blocking Buffer + 0.1% Tween + 0.01% SDS). After three more 5-minute washes in PBS + 0.1 Tween 20, the membranes were imaged using an Odyssey DLx Imaging System (Licor Bioscience).

### HLA-DRB-15*01 imputation and Polygenic Risk Score (PRS) calculation

The tag variant and allele for HLA-DRB-15*01 is rs3135388 and the MS risk allele is “A” (Goris et al. 2008; Zivkovic et al. 2009; Alcina et al. 2012). The genotype of rs3135388 was obtained from whole genome sequence data of each individual. We applied a previously developed and validated PRS (PGS002726) for multiple sclerosis to individuals recruited to this study (Shams et al. 2023). PGS002726 was obtained from the polygenic risk score catalogue (http://www.pgscatalog.org/). We used the nextflow pipeline “The Polygenic Score Catalog Calculator (pgsc-calc)” to calculate each individual’s PRS from whole genome sequencing data (Lambert et al. 2024).

### scRNA-seq

We performed scRNA-seq experiments using cells derived from 5 patients with multiple sclerosis and 5 healthy controls, pooled into one batch. Single-cell 5ʹ immune profiling was performed using the Chromium Next GEM Single Cell 5ʹ Kit v2 (Dual Index, PN-1000263) together with the Single Cell Human BCR Amplification Kit (PN-1000253) from 10x Genomics (Pleasanton, California). Approximately 17,000 cells were loaded onto a Chromium Next GEM Chip K to capture 10,000 cells and processed on a Chromium X Controller to generate barcoded Gel Beads-in-Emulsion (GEMs). GEM-RT, Dynabeads MyOne SILANE cleanup, and cDNA amplification were performed according to the manufacturer’s protocol (CG000331 Rev F). Amplified cDNA was used for both gene expression and BCR V(D)J enrichment and subsequent library construction following the procedures described in the user guide. Final libraries were sequenced on an Illumina NovaSeq 6000 S4 flow cell (PE 100 bp) using the following sequencing parameters: Read 1: 28 cycles; i7 Index: 10 cycles; i5 Index: 10 cycles; Read 2: 90 cycles. Sequencing was performed to a target depth of ∼32,500 read pairs per cell for 5ʹ gene expression libraries and ∼5,000 read pairs per cell for the BCR libraries.

Initial read alignment and quantification of FASTQ files was performed using Cell Ranger (v 6.1.2) (Zheng et al. 2017). Analysis was performed using Seurat (Butler et al. 2018). Cells with less than 1,000 or greater than 7,000 unique features were removed from downstream analysis. No features that were expressed in 3 or fewer cells were included. Data were normalized using the NormalizeData() command. For clustering and Uniform Manifold Approximation and Projection (UMAP) visualization, 15 components were used; this number was determined based on the strength of PCs visualized by elbow plot and PC heatmaps after PCA was performed. The UMAPs and clusters were generated using the FindNeighbors(), FindClusters(), and RunUMAP() functions (R implementation). The DotPlot was created using the DotPlot() function. Input features for the dot plot were generated through the FindAllMarkers() function provided by Seurat to identify differentially expressed immunoglobulin heavy chain variable region (IGHV) reads.

We also analyzed publicly available single cell RNA-seq data. The dataset GSE133028 was downloaded from the Gene Expression Omnibus, consisting of 3 healthy control and 10 relapsing-remitting multiple sclerosis scRNA samples. Raw fastq files for each patient were aligned to the human genome using Cell Ranger (v. 7.0.1) to make a Seurat object. Cells with greater than 15% mitochondrial genes and cells with less than 500 or greater than 4,500 unique features were removed from analysis. All individual Seurat objects were integrated using Harmony (Korsunsky et al. 2019), and B cells were identified using the gene *MS4A1* (otherwise known as CD20) and made into a separate Seurat object. 40 components were used to generate UMAP visualization using the FindNeighbors(), FindClusters(), and RunUMAP() functions. B cell subtypes were identified by comparing the expression levels of select genes based on previously published studies (Verstegen et al. 2023; Dai et al. 2024). *CD21* expression across B cell subtypes was analyzed using the VlnPlot() function in Seurat, and the proportion of CD21+ transitional/naïve B cells were extracted to compare between healthy control and MS. Differential gene expression analysis between healthy control and MS was performed using the FindMarkers() command in Seurat (p-value cutoff = 0.05, logFC cutoff = 0.585).

### ATAC-seq

Transposase Tn5 with adapter sequences was used to cut accessible DNA (Buenrostro et al. 2013). These accessible DNA sequences with adapter sequences were isolated, and libraries were prepared from ∼50,000 primary B cells or EBV-transformed B cells using the OMNI ATAC protocol (Corces et al. 2017). The libraries were sequenced 2x150 bp on an Illumina NovaSeq 6000 at the Cincinnati Children’s Hospital Medical Center (CCHMC) Genomics Sequencing Facility.

ATAC-seq data were processed and aligned to a combined human (hg38) and EBV (B95-8) genome using the ENCODE ATAC-seq pipeline (V2.0.0) (Consortium 2012; Luo et al. 2020; Hitz et al. 2023) (Doi: 10.5281/zenodo.5598324). Aligned reads were then sorted using samtools (v.1.9) and duplicate reads were removed using Picard (v. 2.20.7) (https://broadinstitute.github.io/picard/). Peaks were called within the pipeline using MACS2 (v. 2.2.4) (Zhang et al. 2008). ENCODE’s exclusion list regions were removed from the final set of peaks. Differential chromatin accessibility was calculated using DiffBind (v. 3.0.15) (Stark R 2011), with thresholds of a fold-change greater than 1.5 and adjusted p-value less than 0.05. Each individual-derived cell line was used as a replicate. Expression levels of EBNA2 were statistically controlled for using the DESeq2 design formula “∼log(EBNA2 TPM) + disease_status”.

### ChIP-seq

We performed ChIP-seq in EBV-transformed B cells from 5 MS and 5 HC for EBNA2 (Abcam ab90543; Lot GR3185425-6), RBPJ (Cell Signaling Technology 5313; Lot 3), PU.1 (Cell Signaling Technology 2266; Lot 3), and EBF1 (Santa Cruz sc-137065; Lot J1122) using standard experimental procedures as previously described (Hong et al. 2021). Cells were incubated in a crosslinking solution (1% formaldehyde, 5 mM 4-(2-hydroxyethyl)-1-piperazineethanesulfonic acid (HEPES) pH 8.0, 10 mM sodium chloride, 0.1 mM ethylenediaminetetraacetic acid (EDTA), and 0.05 mM ethylene glycol tetraacetic acid (EGTA)) in Roswell Park Memorial Institute (RPMI) culture medium with 10% fetal bovine serum (FBS) and placed on a tube rotator at room temperature for 10 min. To stop crosslinking, glycine was added to a final concentration of 0.125 M, and the tubes were rotated at room temperature for 5 min. Cells were washed twice with ice-cold phosphate-buffered saline (PBS), resuspended in lysis buffer 1 (50 mM HEPES pH 8.0, 140 mM NaCl, 1 mM EDTA, 10% glycerol, 0.25% Triton X-100, and 0.5% NP-40), and incubated for 10 min on ice. Nuclei were harvested after centrifugation at 5,000 rpm for 10 min, resuspended in lysis buffer 2 (10 mM Tris–HCl pH 8.0, 1 mM EDTA, 200 mM NaCl, and 0.5 mM EGTA), and incubated at room temperature for 10 min. Protease and phosphatase inhibitors (Halt™ Protease and Phosphatase Inhibitor Cocktail (100X), Thermo Fisher Scientific, Waltham, MA) were included in both lysis buffers. Nuclei were resuspended in sonication buffer (10 mM Tris [pH 8.0], 1 mM EDTA, and 0.1% sodium dodecyl sulfate (SDS)). An S220 focused ultrasonicator (COVARIS, Woburn, MA) was used to shear chromatin (150–500-bp fragments) with 10% duty cycle, 175 peak power, and 200 bursts per cycle for 7 min. A portion of the sonicated chromatin was run on an agarose gel to verify fragment sizes. Sheared chromatin was pre-cleared with 10 μL of Protein A or G Dynabeads (Thermo Fisher Scientific) at 4 °C for 1 h.

Immunoprecipitation of TF-chromatin complexes was performed with an SX-8X IP-STAR compact automated system (Diagenode). Beads conjugated to antibodies against each TF were incubated with pre-cleared chromatin at 4 °C for 8 h. The beads were then washed sequentially with wash buffer 1 (10 mM Tris–HCl [pH 7.5], 150 mM NaCl, 1 mM EDTA, 0.1% SDS, 0.1% NaDOC, and 1% Triton X-100), wash buffer 2 (10 mM Tris–HCl [pH 7.6], 400 mM NaCl, 1 mM EDTA, 0.1% SDS, 0.1% NaDOC, and 1% Triton X-100), wash buffer 3 (10 mM Tris–HCl [pH 8.0], 250 mM LiCl, 1 mM EDTA, 0.5% NaDOC, and 0.5% NP-40), and wash buffer 4 (10 mM Tris–HCl [pH 8.0], 1 mM EDTA, and 0.2% Triton X-100). Finally, the beads were resuspended in 10 mM Tris–HCl (pH 7.5) and used to prepare libraries via ChIPmentation.

The ChIP-seq libraries were sequenced as single-end 100-base reads on an Illumina NovaSeq 6000 at the Cincinnati Children’s Hospital Medical Center (CCHMC) Genomics Sequencing Facility. ChIP-seq data were processed and aligned to a combined human (hg38) and EBV (B-958) genome using the ENCODE ChIP-seq pipeline (V2.0.0) (Consortium 2012; Luo et al. 2020; Hitz et al. 2023) (Doi: 10.5281/zenodo.5598352). Adapters were trimmed using fastp (v. 0.23.2) (Chen et al. 2018) prior to processing through the pipeline. Aligned reads were then sorted using samtools (v.1.9) and duplicate reads were removed using Picard (v. 2.20.7) (https://broadinstitute.github.io/picard/). Peaks were called using the pipeline’s default parameters with MACS2 (v. 2.2.4) (Zhang et al. 2008). ENCODE “exclusion list regions” were removed from the final peak set. QC results are available in Supplemental Table S2.

To identify HC- and MS- specific genomic features for ChIP-seq, all peaks that were found in each condition were combined and overlapping events were merged using bedtools (Quinlan and Hall 2010). Peaks unique to HC or MS were identified using the bedtools command “subtract -A”. Shared peaks were determined by removing all condition-specific peaks from the total peak set.

### Enrichment analysis for functional genomic datasets

We used the RELI algorithm to estimate the significance of the overlap between the genomic feature sets (e.g., ChIP-seq peaks) generated in this study (Harley et al. 2018). As input, RELI takes the genomic coordinates of peaks from two datasets. RELI then systematically intersects these coordinates with one another, and the number of input regions overlapping the peaks is counted. Next, a p-value describing the significance of this overlap is estimated using a simulation-based procedure in which the peaks from the first dataset are randomly distributed within the union coordinates of open chromatin from human cells. A distribution of expected overlap values is then created from 2,000 iterations of randomly sampling from the negative set, each time choosing a set of negative examples that match the input set in terms of the total number of genomic loci. The distribution of the expected overlap values from the randomized data resembles a normal distribution and can thus be used to generate a Z-score and corresponding p-value estimating the significance of the observed number of input regions that overlap each dataset.

### GWAS data set curation

We obtained GWAS data from the NHGRI-EBI GWAS catalog (Buniello et al. 2019) and the supplemental dataset from the International Multiple Sclerosis Genetics Consortium GWAS publication (International Multiple Sclerosis Genetics 2019). A genome-wide significance cutoff of 5 × 10^−8^ was used to establish the statistical significance of a variant and its association to a given disease or trait. Independent loci were identified using LD-based pruning in PLINK (Purcell et al. 2007) (window size 300,000 kb, SNP shift size 100,000 kb, and *r*^2^ < 0.2).

### Pathway enrichment analysis

Pathway analysis for MS and HC differentially expressed genes was performed using Enrichr (Chen et al. 2013; Kuleshov et al. 2016).

### TF DNA binding motif enrichment analysis

The HOMER software package (v. 4.9) (Heinz et al. 2010) was used to calculate TF DNA binding site motif enrichment for a custom library containing the human CisBP database motifs (build 2.0) (Weirauch et al. 2014; Lambert et al. 2019). HOMER was modified to use a log_2_-based likelihood scoring system.

### Identification of allele-dependent sequencing reads using MARIO

Allele-dependent behavior was identified in sequencing reads using the MARIO pipeline (Harley et al. 2018). Briefly, the MARIO pipeline identifies allele-dependent behavior by weighing (1) the imbalance between the number of reads that are mapped to each allele, (2) the total number of reads mapped at each variant, and (3) the number and consistency of available replicates. These variables are combined into a single Allelic Reproducibility Score (ARS), which reflects the degree of allelic behavior observed for the given heterozygous variant in the given data set. MARIO ARS values exceeding 0.4 were considered to be allelic, following our previous study (Harley et al. 2018). We implemented the MARIO pipeline for EBNA2 ChIP-seq data from this study along with publicly available EBNA2 binding data (McClellan et al. 2013; Hong et al. 2021). To select for consistent allele-dependent binding activity, we identified and graphed ChIP-seq peaks with at least 9 reads on top of an MS variant and with at least 15% difference between the reads mapping to the strong and weak alleles (Fig. 5A, Supplemental Table S8).

To expand our reach, we assessed additional EBNA2 ChIP-seq experiments performed in our laboratory, including the cell lines GM12878, IB4, AG876, and Jiyoye. Additionally, we included MS risk variants with published allelic EBNA2 binding activity (Supplemental Table S8) (Harley et al. 2018; Hong et al. 2021). All identified MS risk variants that were overlapped by EBNA2 ChIP-seq peaks in an allelic manner were assessed for eQTL function using release 7 of the eQTL Catalogue (Kerimov et al. 2021). Gene targets of these eQTLs (p-value < 0.01) were used as input to STRING (Szklarczyk et al. 2023) (Fig. 5C).

## Data Availability

All data produced in the present work are contained in the manuscript

## Acknowledgements

This research was funded by the National Institutes of Health (NIH) R01 NS099068, P30 AR070549, R01 AR073228, and R01 AI024717 to M.T.W. and L.C.K.; U19 AI070235, R01 DK107502, R01 AI148276, and U01 HG011172 to L.C.K.; R01 HG010730, R01 GM055479, U01 AI130830, U01 AI150748, R01 AI141569, P01 AI150585, and U24 HG013078 to M.T.W. We thank the Single Cell Genomics Core (RRID:SCR_022653) at the Cincinnati Children’s Hospital Medical Center, especially Kelly Rangel, for their assistance with scRNA-seq. We thank the Cincinnati Children’s Hospital Medical Center (CCHMC) Genomics Sequencing Facility (RRID:SCR_022630). We thank John Harley for helpful discussions around the role of EBNA2 in autoimmunity.

## Author contributions

Marissa Granitto and Ellie Kim contributed equally to this work. Matthew Weirauch and Leah Kottyan co-led this work. Conceptualization: M.G., L.C.K., M.T.W.; Study design: M.G., L.C.K., M.T.W., C.F.; Participant recruitment: C.F., V.B., K.F., A.Z.; Statistical analysis design: M.G., L.C.K, M.T.W.; Computational analysis: M.G., P.J.D., A.V.H. S.P., E.K., C.O.S., X.C., L.E.E., K.K., K.F., B.G.; Directed experiments: M.G., C.F., L.C.K., M.T.W.; Performed Experiments: M.G., C.F., C.Y., M.S.S., O.D., A.A.D., K.D., V.L., D.H.; Writing: M.G., E.K., L.C.K., M.T.W, L.P.L.; Funding: L.C.K., M.T.W. All authors reviewed and approved the final manuscript.

