## Supplemental Figure S1 for "Enhanced EBNA2-dependent activity in EBV-transformed B cells from patients with multiple sclerosis"

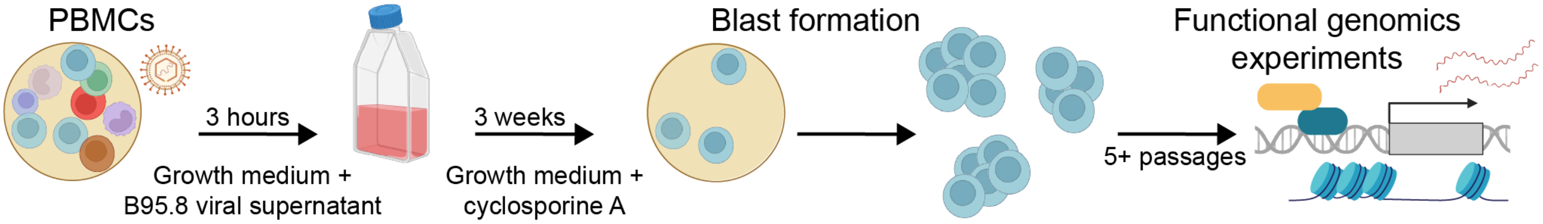


**Figure S1. EBV-transformed B cell line generation.** Schematic depicts the method used to make EBV-transformed B cell lines from peripheral blood mononuclear cells (PBMCs) derived from individuals who are healthy controls or have multiple sclerosis. See Methods.
