## Supplemental Figure S2 for "Enhanced EBNA2-dependent activity in EBV-transformed B cells from patients with multiple sclerosis"

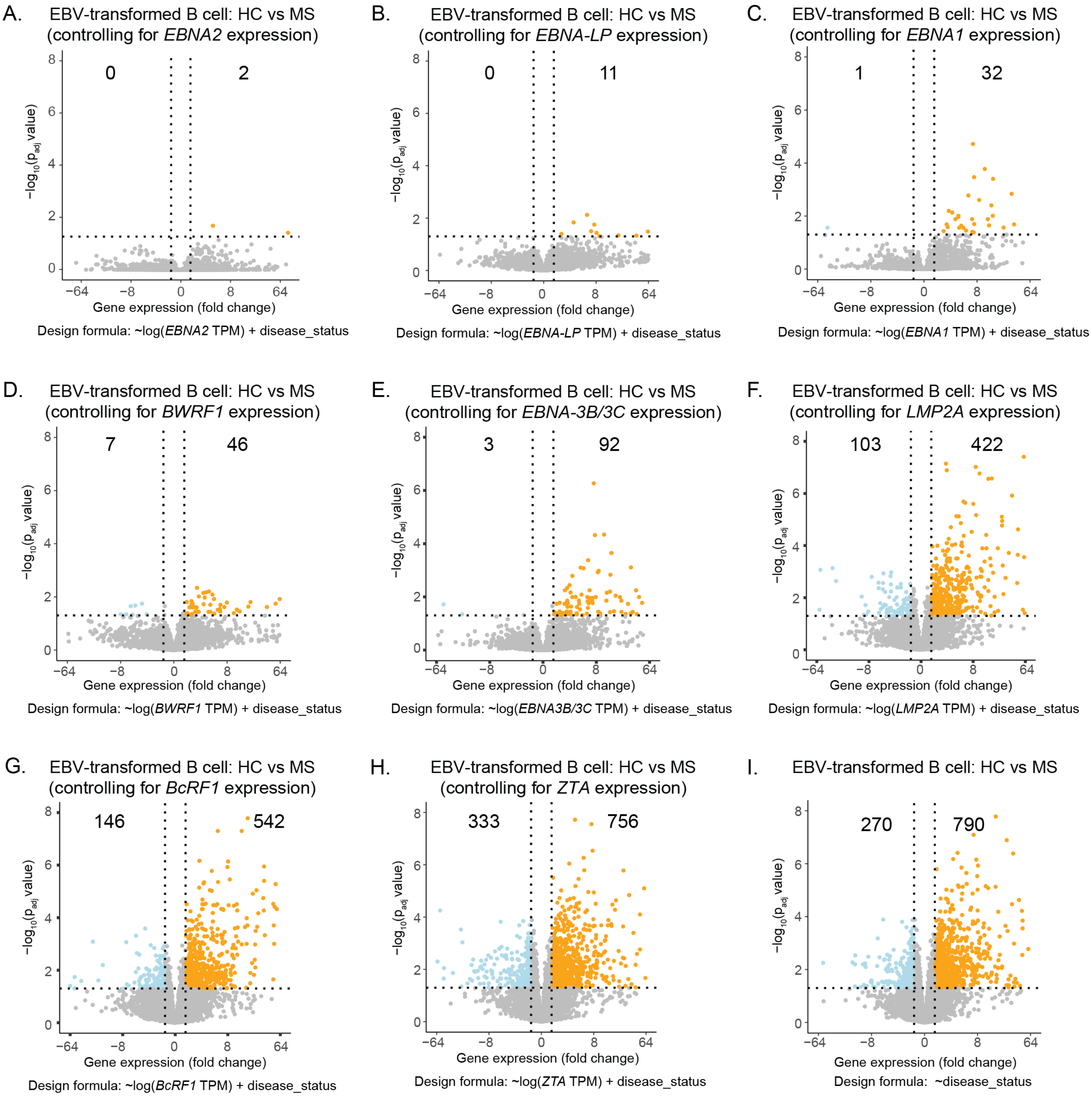


**Figure S2. EBNA2 expression level accounts for the largest amount of differentially expressed genes compared to other EBV genes.** Comparison of human gene expression levels (RNA-seq) in healthy controls (HC) and patients with multiple sclerosis (MS). Horizontal dashed lines indicate an adjusted p-value significance threshold of 0.05. Vertical dashed lines indicate a fold-change threshold of 1.5. Numbers indicate gene counts. The design formula used in DESeq2 is provided under each panel. A-H. Results for EBV- transformed B cells after statistically adjusting for the expression levels of the indicated gene. All differentially expressed EBV genes and one additional lytic gene (ZTA) are included. I. Results for EBV- transformed B cells without adjustment. Panel A is also shown in Figure 1C. Panel I is also shown in Figure 1B.
