## Supplemental Figure S3 for "Enhanced EBNA2-dependent activity in EBV-transformed B cells from patients with multiple sclerosis"

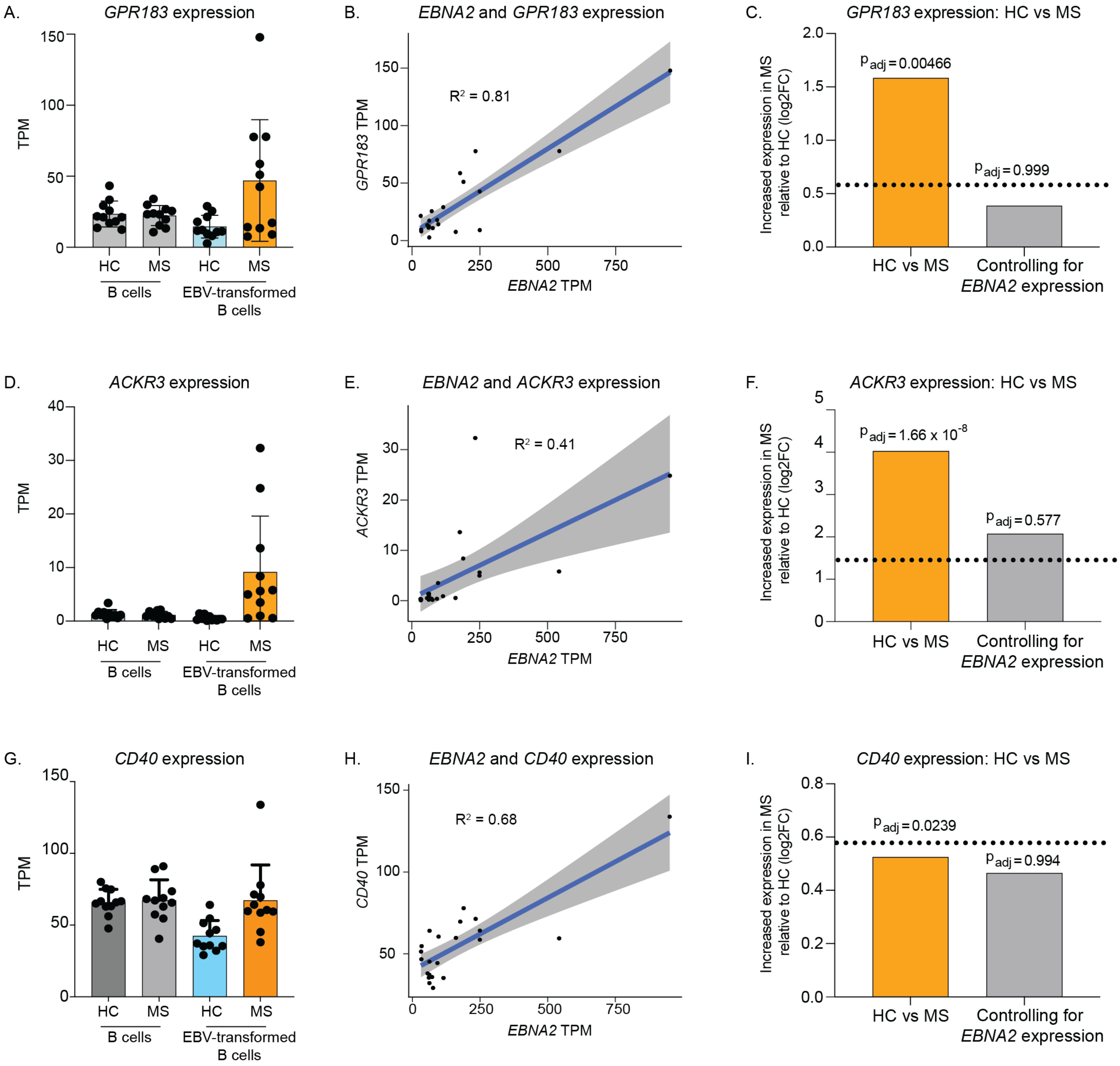


**Figure S3. EBNA2-dependent changes in gene expression.** A. Gene expression of GPR183 (RNA-seq) is shown in transcripts per million (TPM). Each dot represents a cell line derived from one individual. Error bars indicate standard deviation. B. Correlation of EBNA2 and GPR183 expression (TPM). C. GPR183 differential expression as a function of HC vs MS with and without controlling for EBNA2 expression. Dashed line indicates a fold-change of 1.5. D. Gene expression of ACKR3 (RNA-seq) is shown in transcripts per million (TPM). Each dot represents a cell line derived from one individual. Error bars indicate standard deviation. E. Correlation of EBNA2 and ACKR3 expression (TPM). F. ACKR3 differential expression as a function of HC vs MS with and without controlling for EBNA2 expression. Dashed line indicates a fold-change of 1.5. G. Gene expression of CD40 (RNA-seq) is shown in transcripts per million (TPM). Each dot represents a cell line derived from one individual. Error bars indicate standard deviation. H. Correlation of EBNA2 and CD40 expression (TPM). I. CD40 differential expression as a function of HC vs MS with and without controlling for EBNA2 expression. Dashed line indicates a fold-change of 1.5.
