## Supplemental Figure S4 for "Enhanced EBNA2-dependent activity in EBV-transformed B cells from patients with multiple sclerosis"

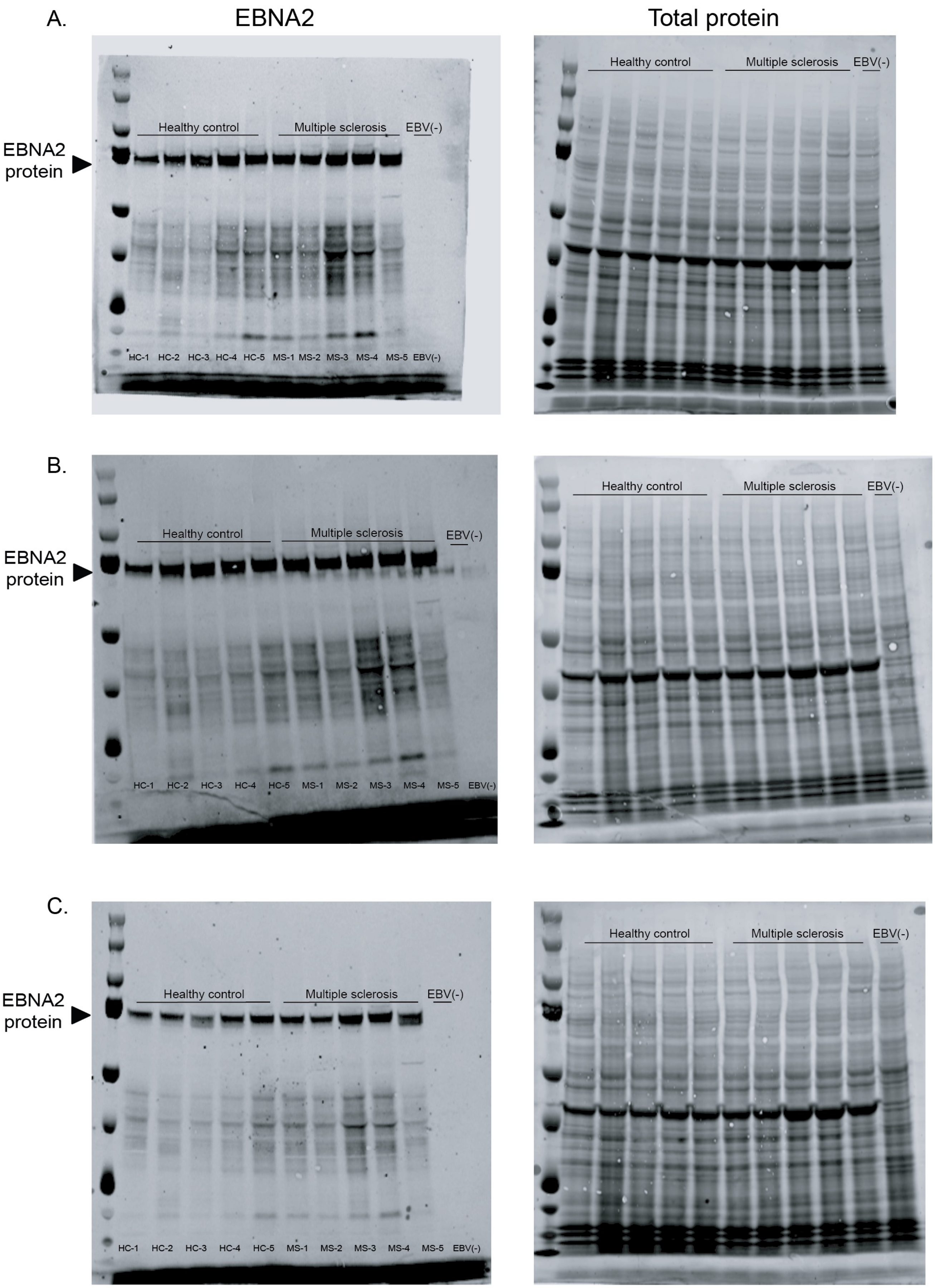


**Figure S4. Increased EBNA2 protein in MS-derived EBV-transformed B cells**. A-C. Three replicates of Western blots for EBNA2 protein (left) and total protein stain (right) for 5 HC and 5 MS EBV-transformed B cell lines and an Akata EBV-negative B cell line. The order of the lanes is as follows: ladder (1), HC (2-6), MS (7-11), EBV-negative B cell line (12).
