## Supplemental Figure S5 for "Enhanced EBNA2-dependent activity in EBV-transformed B cells from patients with multiple sclerosis"

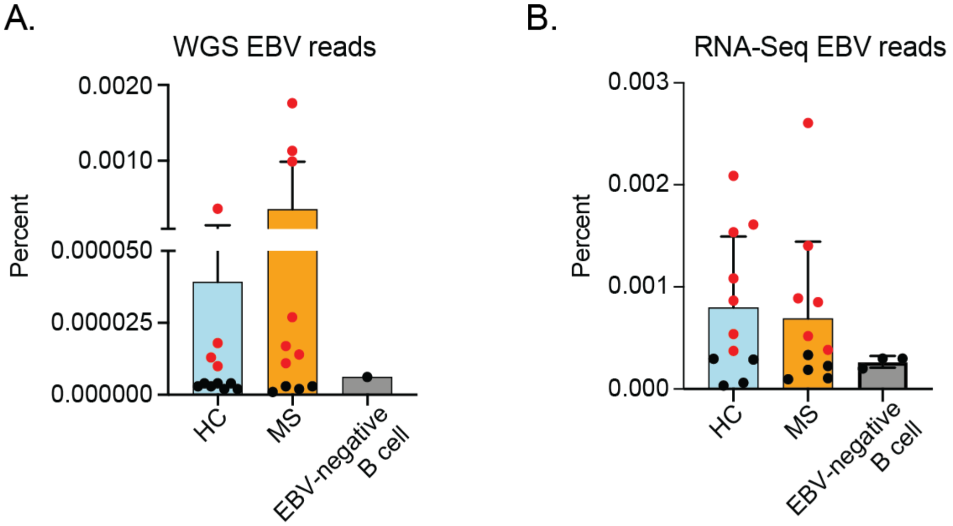


**Figure S5. Evidence of prior exposure of individuals in this study to EBV.** A. The total percentage of EBV-mapped whole genome sequencing reads from PBMCs derived from HC and MS, and an EBV-negative B cell line (Akata negative). Each dot represents DNA from PBMCs derived from one individual. Error bars indicate standard deviation. Dots colored red were designated as indicators of prior EBV exposure to compliment ELISA assays reported in Additional file 1: Table S1. B. The total percent of RNA sequencing reads mapped to the EBV genome in primary B cells from HC and MS. An EBV-negative B cell line was used as a negative control. Each dot indicates one individual. Error bars indicate standard deviation. Dots colored red were designated as indicators of prior EBV exposure to compliment ELISA assays reported in Additional file 1: Table S1.
