## Supplemental Figure S6 for "Enhanced EBNA2-dependent activity in EBV-transformed B cells from patients with multiple sclerosis"

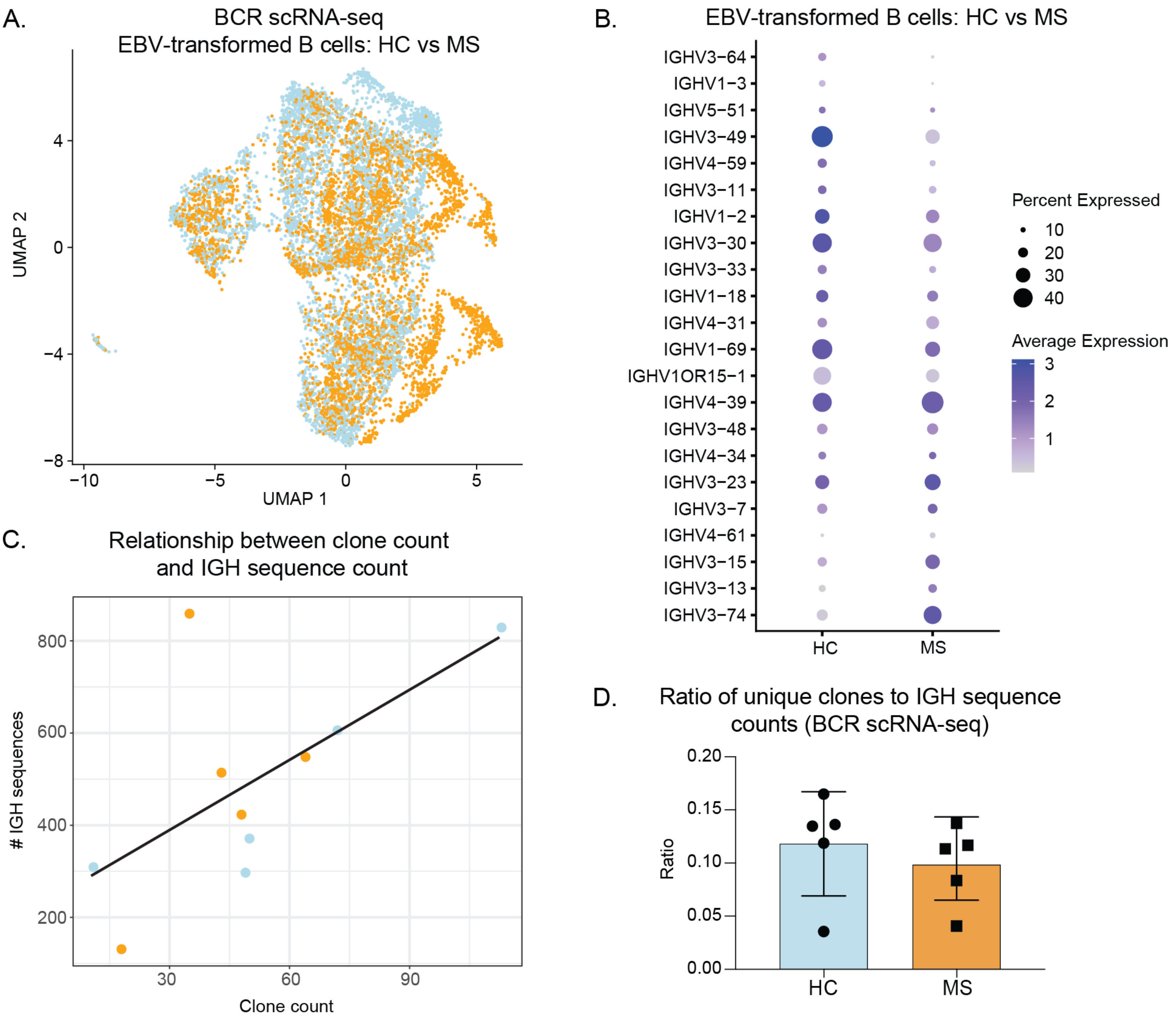


**Figure S6. Single cell RNA-sequencing with BCR enrichment allows for enumeration of unique clones foreach EBV-transformed B cell line.** A. UMAP depicting EBV-transformed B cells passing quality filters (see Methods).Cells derived from 5 healthy controls (HC, blue) and 5patients with multiple sclerosis (MS, orange) are shown. B. Comparison of pseudobulk human immunoglobulin gene expression levels (scRNA-seq) in HC and MS. The B cell receptor (also known as BCR or immunoglobulin) contains heavy and light chains. The genes encoding the heavy and light chains include sequences that are uniquely rearranged during B cell selection. Dot plot of all IGHV genes ordered by fold-change. C. Relationship between clone count and IGH sequence count. Single cell data are sparse (i.e., not all B cells expressing immunoglobulin will have single cell RNA sequencing reads to immunoglobulin genes). We expect the number of cells with detected immunoglobulin will influence the number of clones detected. The number of unique clones for each cell line was determined based on the number of unique immunoglobulin heavy chain (IGH) sequences. The total number of IGH sequences was used to approximate the number of B cells. Each dot represents the number of unique clones (x axis) graphed as a function of number of total IGH sequences (y axis) for one individual. The trend line indicates the expected correlation between the number of IGH sequences and clone count. D. Based on the strong positive correlation between the number of IGH sequences and clone count, we also calculated the ratio of unique clones to IGH sequences for each sample (y axis) as a function of disease status (x axis).
