## Supplemental Figure S7 for "Enhanced EBNA2-dependent activity in EBV-transformed B cells from patients with multiple sclerosis"

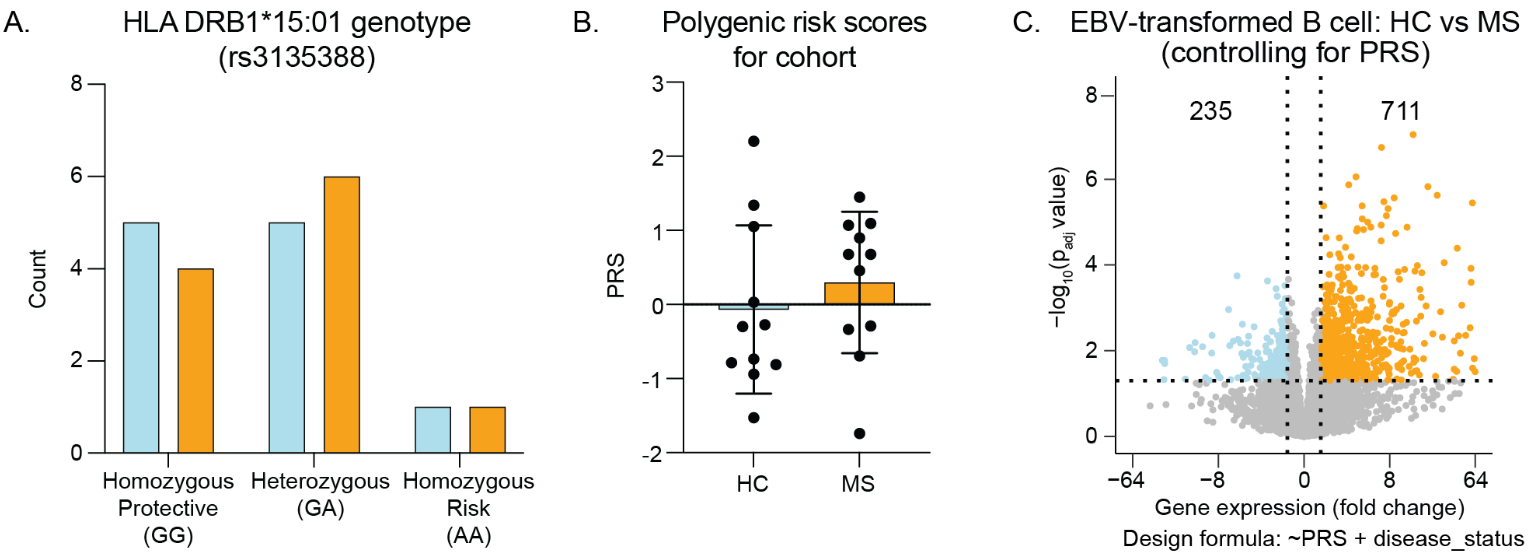


**Figure S7. Multiple sclerosis genetic risk variants do not fully account for differential gene expression in EBV-transformed B cells derived from HC and MS.** A. The number of healthy controls (HC, blue) and patients with multiple sclerosis (MS, orange) with each HLA-DRB1*15:01 genotype. The genetic variant alleles of rs3135388 result in the differential amino acid use that allows for imputation of the HLA type for each individual. B. Polygenic risk scores (PRS) were calculated for each individual based on PGS002726 [56]. See Methods. Each dot indicates one individual. Error bars indicate standard deviation. C. Comparison of human gene expression levels (RNA-seq) in HC and MS after statistically adjusting for PRS. Horizontal dashed lines indicate an adjusted p-value significance threshold of 0.05. Vertical dashed lines indicate a fold-change threshold of 1.5. Numbers indicate gene counts. The design formula used in DESeq2 is provided under the panel.
