## Supplemental Figure S8 for "Enhanced EBNA2-dependent activity in EBV-transformed B cells from patients with multiple sclerosis"

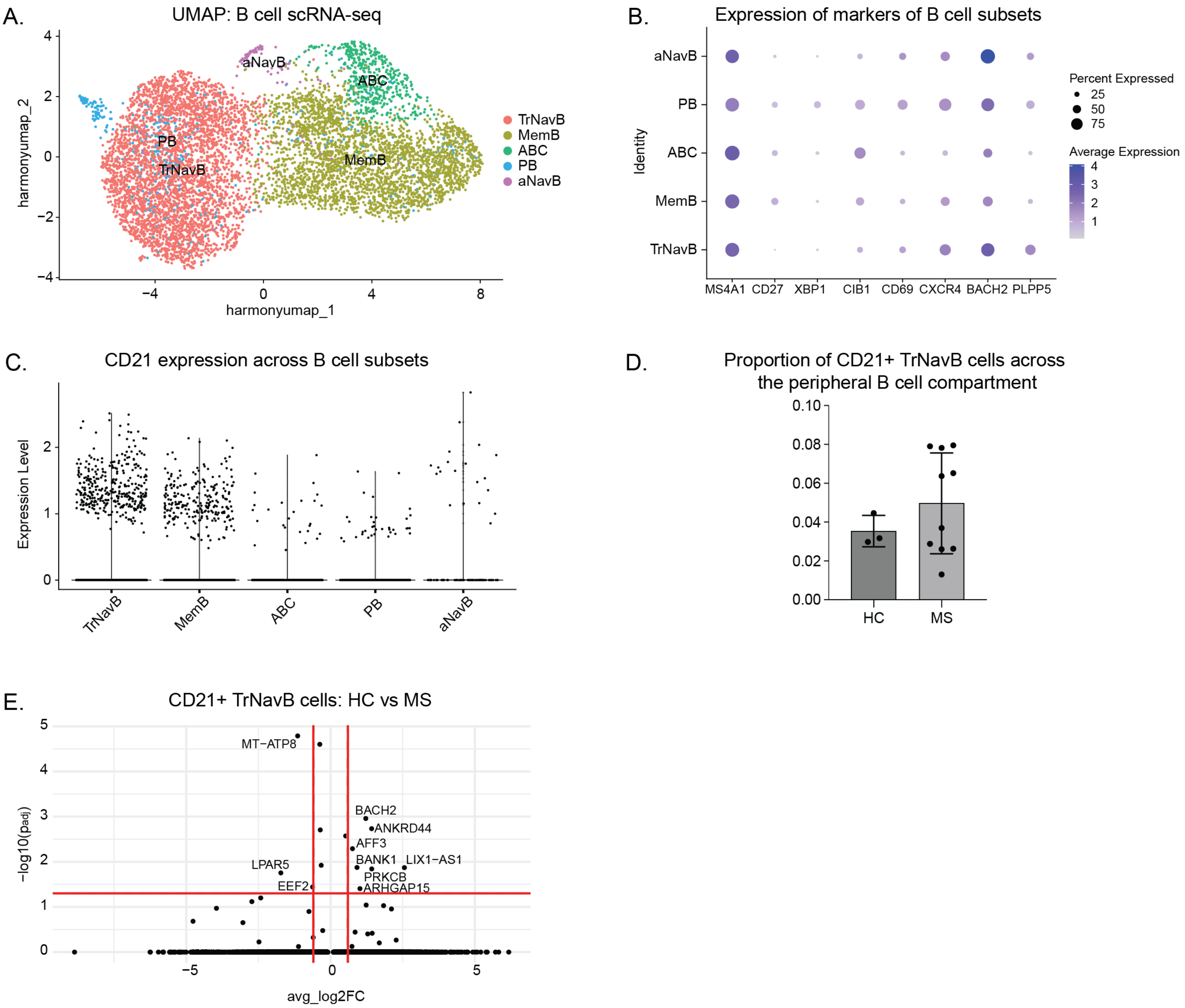


**Figure S8. There are few transcriptional differences in CD21+ transitional/naïve B cells between HC and MS**. A. UMAP with labelled B cell subsets across all patients (n=13) from GSE133028 cohort. B. Dotplot of genes used to label B cell subsets from A. Prior to identifying cell type, PCA analysis yielded 7 clusters (0 through 6). Clusters 0 and 3 were identified as Transitional/Naïve by referencing the paper by Dai, et al, 2024. Cluster 4 was identified as “age related B cell” (ABC) based on the high expression of CIB1. Cluster 6 was identified as activated NavB based on its similar transcriptional profile to cluster 4. Cluster 5 was identified as a plasmablast/cell based on the expression of XBP1 (Verstegen, 2023). Clusters 1 and 2 were identified as MemB cells based on the relatively high expression of CD27 (Dai, 2024 and Verstegen, 2023). C. Violin plot of CR2 (CD21) expression across B cell subsets. D. Bar graph of the proportion of CD21+ TrNav B cells in HC and MS. E. Comparison of human gene expression levels in HC and MS CD21+ TrNavB cells. Horizontal red lines indicate an adjusted p-value significance threshold of 0.05. Vertical red lines indicate a fold-change threshold of 1.5.
